# Single-cell atlas of transcriptomic vulnerability across multiple neurodegenerative and neuropsychiatric diseases

**DOI:** 10.1101/2024.10.31.24316513

**Authors:** Donghoon Lee, Mikaela Koutrouli, Nicolas Y. Masse, Gabriel E. Hoffman, Seon Kinrot, Xinyi Wang, N.M. Prashant, Milos Pjanic, Tereza Clarence, Fotios Tsetsos, Deepika Mathur, David Burstein, Karen Therrien, Aram Hong, Clara Casey, Zhiping Shao, Marcela Alvia, Stathis Argyriou, Jennifer Monteiro Fortes, Pavel Katsel, Pavan K. Auluck, Lisa L. Barnes, Stefano Marenco, David A. Bennett, PsychAD Consortium, Lars Juhl Jensen, Kiran Girdhar, Georgios Voloudakis, Vahram Haroutunian, Jaroslav Bendl, John F. Fullard, Panos Roussos

**Author notes:** These authors contributed equally to this work (Donghoon Lee, Mikaela Koutrouli, Nicolas Y. Masse, Gabriel E. Hoffman, John F. Fullard, Panos Roussos).

## Abstract

Neurodegenerative and neuropsychiatric diseases impose a significant societal and public health burden. However, our understanding of the molecular mechanisms underlying these highly complex conditions remains limited. To gain deeper insights into the etiology of different brain diseases, we used specimens from 1,494 unique donors to generate a population-scale single-cell transcriptomic atlas of the human dorsolateral prefrontal cortex (DLPFC), comprising over 6.3 million individual nuclei. The cohort includes neurotypical controls as well as donors affected by eight common and complex brain disorders: Alzheimer’s disease (AD), diffuse Lewy body disease (DLBD), vascular dementia (Vas), Parkinson’s disease (PD), tauopathy, frontotemporal dementia, schizophrenia, and bipolar disorder. We show that inter-individual variation accounts for a substantial portion of gene expression variation in the DLPFC. By comparing transcriptomic variation across diseases, we reveal universal signatures enriched in basic cellular functions such as mRNA splicing and protein localization. After discounting these cross-disease signatures, we show strong genetic and transcriptomic concordance among AD, DLBD, Vas, and PD, largely driven by alteration of synaptic signaling functions in neurons. Furthermore, we characterize transcriptomic variation among different AD phenotypes that were distinct from healthy aging. We uncover mitigating effects of interneurons and aggravating effects of immune and vascular cells in AD dementia. Further exploring the effect of the neuropsychiatric symptoms frequently accompanying AD, we identify a link to deep layer excitatory neurons. By constructing transcriptome trajectories that capture AD progression, we show cell-type specific responses implicated in early and late stages of AD. Our atlas provides an unprecedented perspective of the transcriptomic landscape in neurodegenerative and neuropsychiatric diseases, shedding light on shared and distinct processes involving the neuro-immune-vascular systems, and identifying potential targets for therapeutic intervention.

## Main

The human brain is a highly complex organ composed of billions of functionally diverse cells. Under pathogenic stress, cellular and molecular responses are often convoluted and contextual, so understanding their dysfunction in disease is challenging. Recent work has begun to unravel the molecular changes that occur at the single-cell level, which has been particularly helpful in understanding the vulnerability of specific cell types in various disease contexts, as well as the complex interplay between different cell types. In Alzheimer’s disease (AD), it has been shown that a thorough exposition of cellular heterogeneity in the brain, the coordinated interactions between neurons and glia, and the selective depletion of vulnerable inhibitory neuronal subtypes are critical for understanding AD pathology^1,2^.

Building a large-scale disease atlas at single-cell resolution creates an unprecedented opportunity to understand molecular responses at the cellular level and estimate population-level variation in the brain transcriptome. A large sample size provides the resolution needed to establish robust basal-level conditions and to sufficiently capture the full spectrum of disease pathology. By leveraging cross-disease atlases, studies have revealed shared and distinct patterns of gene-expression perturbations in major psychiatric diseases as well as shared genetic factors leading to molecular convergence^3,4^. As such, characterizing shared transcriptomic vulnerabilities and pathophysiology together has significant implications for early treatment and the development of effective therapeutics.

Here, we introduce the PsychAD cohort, which consists of 1,494 unique brain donors affected by various neurodegenerative and neuropsychiatric diseases in addition to neurotypical controls. The resulting single-nucleus RNA sequencing (snRNA-seq) dataset in the dorsolateral prefrontal cortex (DLPFC) of those donors, comprising over 6.3 million nuclei representing 27 distinct subclasses of cells, is sufficiently-powered to identify molecular signatures from multiple traits while accounting for individual variation. We utilize this dataset to uncover shared transcriptomic vulnerability across these diseases and, in so doing, to better understand specific transcriptional patterns and regulatory drivers underpinning each trait. By characterizing putative disease-driving molecular changes across multiple traits, we differentiate shared and novel disease- and cell type-specific associations. Concordance between heritability estimates and transcriptomic similarity identifies shared genetic factors underlying cross-disorder traits. Deep phenotyping of AD trajectories using Tau pathology and clinical dementia status suggests a potential link between immune and brain vasculature dysfunctions. In summary, our study provides a rich and comprehensive resource for exploring the cellular and molecular mechanisms of brain function and dysfunction across multiple neurodegenerative and neuropsychiatric diseases.

### The PsychAD cohort represents diverse neurodegenerative and neuropsychiatric diseases across the lifespan

The PsychAD cohort comprises 1,494 unique brain donors (**Fig. 1a, Supplementary Table 1**). Brain tissue specimens were obtained from three sources: 1,042 donors from Mount Sinai NIH Neurobiobank (MSSM), 300 from Human Brain Collection Core (HBCC), and 152 from Rush Alzheimer’s Disease Center (RADC). The cohort covers the whole lifespan of postnatal ages between 0 and 108, roughly equal numbers of males and females, and represents a diverse range of disease phenotypes, including Alzheimer’s disease (AD), diffuse Lewy body disease (DLBD), vascular dementia (Vas), tauopathy (Tau), frontotemporal dementia (FTD), Parkinson’s disease (PD), schizophrenia (SCZ), and bipolar disorder (BD). The cohort covers a diverse genetic background and over 30% of the donors were of non-European (EUR) ancestry.

**Fig. 1.**
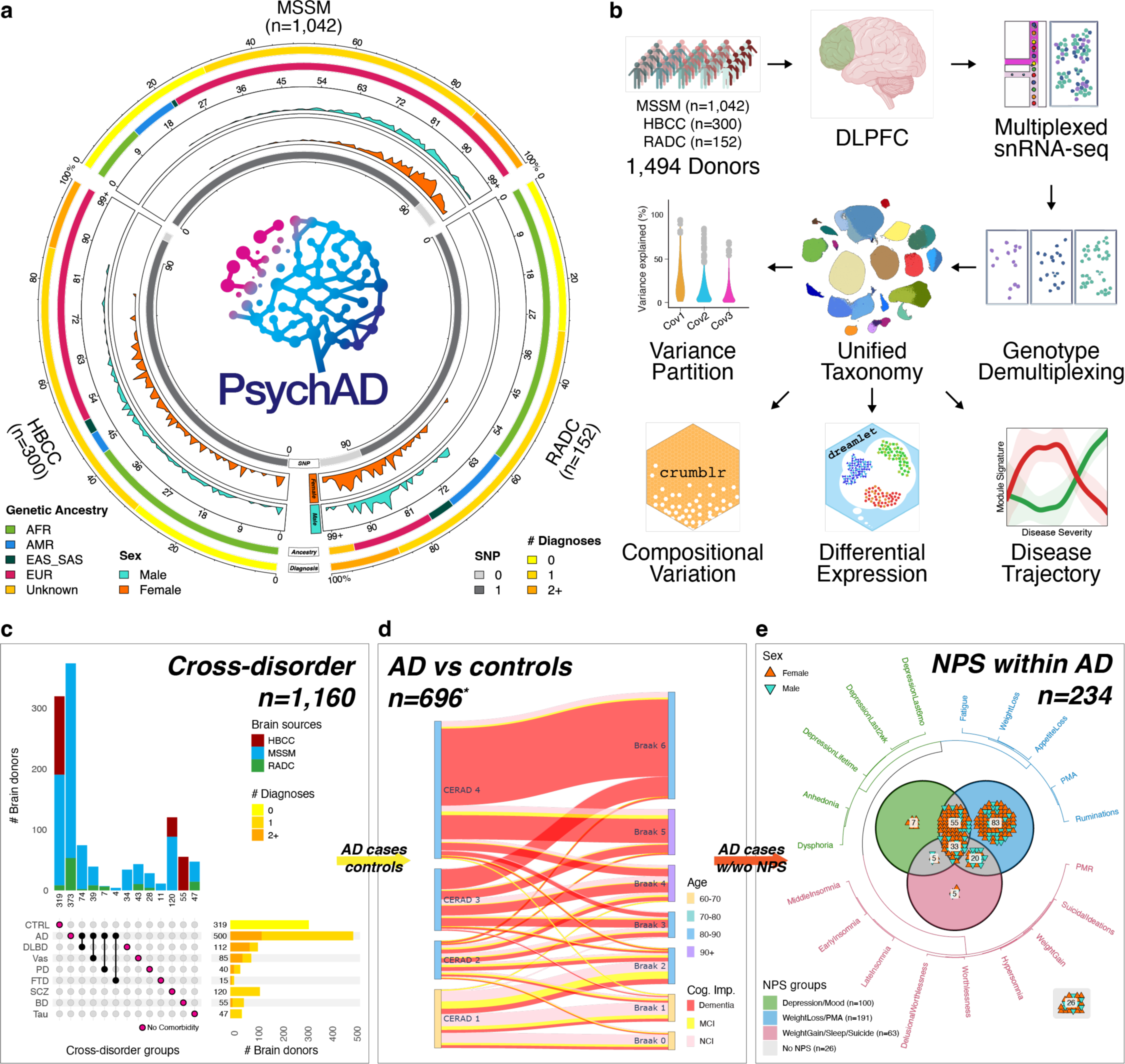
Overview of the PsychAD cohort and study design. **(a)** Breakdown of donors by tissue source, number of diagnoses, genetic ancestry, distribution of age at death, sex, and % availability of genotype data. **(b)** Data generation and analysis workflow. **(c)** A subset of the PsychAD cohort (n=1,160 donors) focused on cross-disorder contrasts. This subset includes a group of neurotypical controls that were used to compare against 8 neurodegenerative and neuropsychiatric disorders. UpSet plot describing available disease phenotypes and number of unique donors by brain bank. **(d)** A subset of the PsychAD cohort (n=696 donors) focused on AD phenotype contrasts. Comparison of different measures of AD severity, including neuritic plaque density (CERAD), neurofibrillary tangle (NFT) pathology, and cognitive impairments. *n=54 donors with incomplete pathology or cognitive status were omitted from the figure. **(e)** A subset of the PsychAD cohort (n=234 donors) was used to characterize single-cell transcriptomic changes underlying NPS in AD. AFR: African, AMR: Ad Mixed American, EAS: East Asian, SAS: South Asian, EUR: European, NPS: Neuropsychiatric Symptom, PMA: Psychomotor Agitation, PMR: Psychomotor Retardation.

We streamlined the unified processing of the data as well as the harmonization of clinical, and technical metadata (**Fig. 1b, Supplementary Fig. 1a, Methods**). Frozen brain specimens were randomized and processed in batches of 6. Equal numbers of nuclei from each sample were pooled together and each pool was subjected to snRNA-seq twice to generate a technical replicate. Following quality control (**Supplementary Figs. 1c-i**), the final data sets consisted of 6,320,459 nuclei. To characterize transcriptomic vulnerability across multiple neuropsychiatric and neurodegenerative diseases, we organized the analysis into three tiers.

First, we focused on the cross-disorder analyses (**Fig. 1c**). We targeted six neurodegenerative diseases (NDDs, including AD, DLBD, Vas, Tau, FTD, and PD) and two neuropsychiatric diseases (NPDs, including SCZ and BD), using a subset of 1,160 donors with minimal comorbidity aged ≥ 17. To estimate the sharing of transcriptomic vulnerability, we compared donors affected with NDDs and NPDs against the baseline of 319 neurotypical controls.

The second-tier analysis focused on the stage of AD progression based on pathological and cognitive impairment measures using a subset of 696 individuals (**Fig. 1d**). To uncover cell-type-specific roles in disease onset and disease trajectory, we compared two characteristic neuropathological abnormalities of the AD brain: the accumulation of amyloid-β (Aβ) plaques measured using CERAD plaque density score and tau-based neurofibrillary tangle (NFT) pathology measured using Braak staging, along with cognitive impairment.

In the third tier, we surveyed neuropsychiatric symptoms (NPS) within 234 individuals affected by AD pathology (**Fig. 1e, Supplementary Fig. 1b**). NPS are core features of AD and are common in patients with dementia^5^. We broadly categorized NPS into three groups, (1) depression or mood-related, (2) weight loss or psychomotor agitation (PMA), and (3) weight gain, insomnia, or suicidal ideation, based on co-occurrence estimates, sharing molecular mechanisms that lead to increased prevalence with disease severity.

### Unified processing and hierarchical cellular taxonomy of the human prefrontal cortex uncovers 27 distinct subclasses of cells

To understand heterogeneous human cortical tissues in disease contexts, we require a cell type taxonomy that is robust to aging, disease phenotypes, and various sampling and technical biases. Following unified computational processing, quality controls, and batch normalization of snRNA-seq libraries representing 1,494 dissections, processed in duplicate (**Methods**), we annotated cell types of the human DLPFC using the cell taxonomy of the primate DLPFC^6^ and human primary motor cortex^7^ as a baseline reference. The resulting human DLPFC cellular taxonomy was organized using three levels of hierarchy, identifying 8 broad cell classes, 27 subclasses, and 65 functionally distinct subtypes (**Fig. 2a, Supplementary Table 2**). Each level of the annotation hierarchy represents a slice in the clustering dendrogram. At the top, the “class level” of the annotation hierarchy defines 8 major cell types, including two broad neuronal cell types: glutamatergic excitatory (EN) and GABAergic inhibitory neurons (IN), three glial: astrocytes (Astro), oligodendrocytes (Oligo), and oligodendrocyte progenitor cells (OPC), and three non-neuronal cell types: immune cells (Immune), mural and vascular cells (Mural), and endothelial cells (Endo). Subsequent levels of the annotation hierarchy, subclasses and subtypes, were derived by iteratively re-clustering the subset of cells by gene matrix using a new set of variable genes relevant to the particular cell type (see *iterative clustering* in **Methods**). The subclass level distinguished the EN class into 10 subclasses and the IN class into 7 subclasses (**Supplementary Figs. 2c,d**). Different types of neurons, especially ENs, are organized into six horizontal layers (L1-L6) that are distinct in both cytoarchitecture and function^8^. *In situ* spatial transcriptomics data was used to confirm that the EN subclasses were spatially distinct and found in their respective neocortical layers (**Fig. 2b**). The EN subclasses were denoted by their laminar organization (L2-6) and axon projection characteristics (IT: intra-telencephalic, ET: extra-telencephalic, NP: near projecting, CT: corticothalamic, and L6B). IN subclasses were determined using their characteristic marker genes (**Figs. 2c,d**); Ivy cells (IN_LAMP5_LHX6), neurogliaform cells (IN_LAMP5_RELN), basket cells (IN_PVALB), chandelier cells (IN_PVALB_CHC), Martinotti and non-Martinotti cells (IN_SST), VIP (IN_VIP), and homologs of mouse Sncg inhibitory neurons^6^ (IN_ADARB2). Unlike laminar organization of EN, IN subclasses were distributed randomly throughout the gray matter, except for IN_ADARB2, which was predominantly found in the superficial layer of the neocortex (**Supplementary Fig. 2e**). Some previously annotated rare inhibitory neuron types, like SST NPY or SST HGF, were not distinguished at the subclass level, but were identified at the subtype level. The cellular taxonomy was relatively consistent, and the subtypes were well represented across all three brain sources (**Supplementary Figs. 2a,b**). Neuronal cells made up 38.4% (EN 23.0% and IN 15.4%), with oligodendrocytes being the next most abundant, at 36.1%. Major cell types matched well when compared to the previous cellular taxonomy of DLPFC^1,6^, and subtypes were relatively concordant (**Supplementary Figs. 2f-h**). Neuronal subclasses exhibit distinct functional characteristics from non-neuronal subclasses (**Fig. 2e**). ENs are enriched in pathways associated with synaptic vesicle priming and neurotransmitter secretion, whereas INs are enriched with functions related to receptor signaling and ion transport. Immune cell types show unique enrichments in functions related to cytotoxic immune responses, while mural cells are involved in vessel morphogenesis. It has been shown cell types are differentially implicated in mediating disease risks^9,10^. Therefore, we further annotated cell types with disease GWASs using the single-cell disease-relevance score (scDRS) and found that neurological diseases largely involve immune and glial cell types, whereas psychiatric diseases are predominantly associated with neuronal cell types (**Fig. 2f, Supplementary Fig. 2i**).

**Fig. 2.**
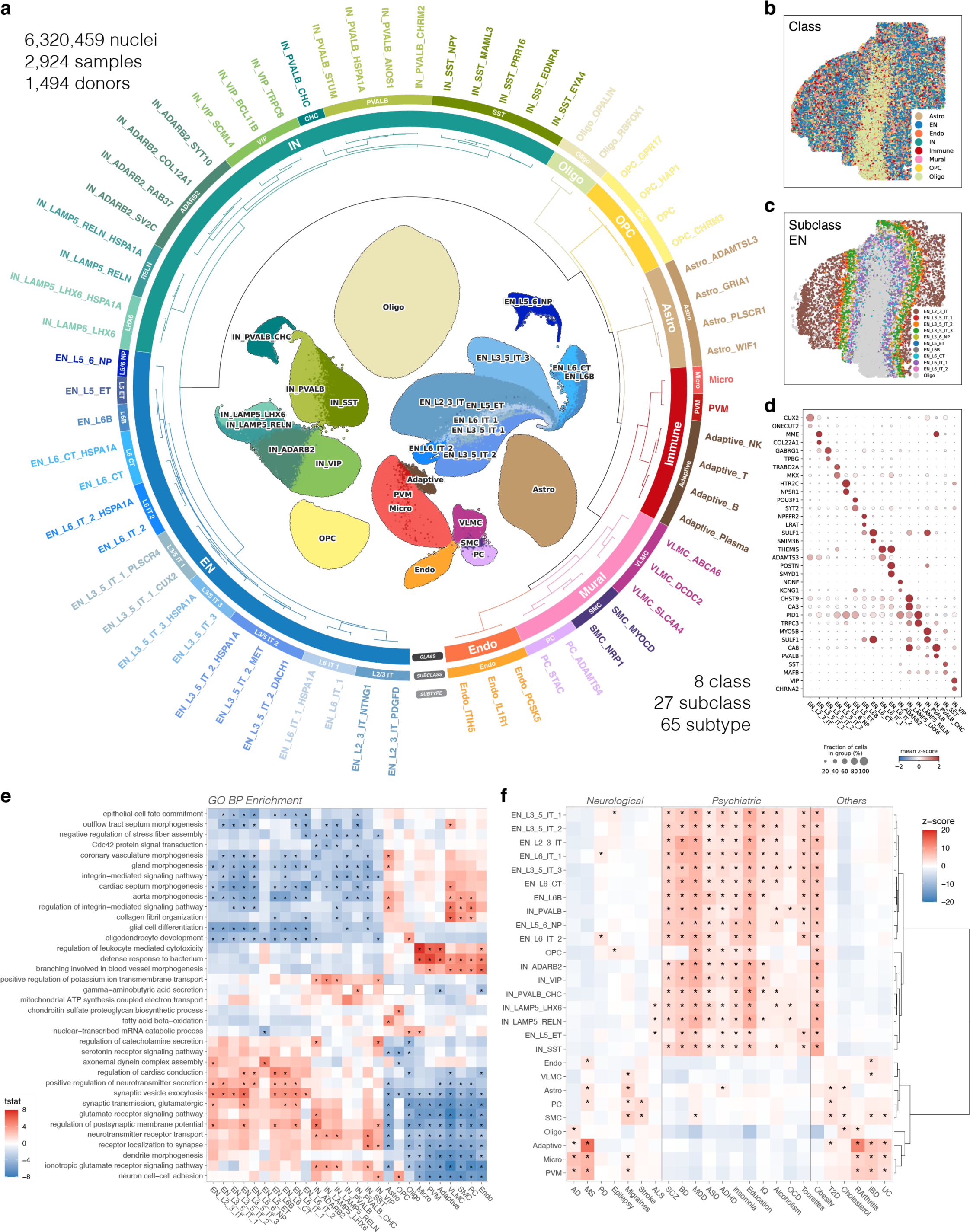
Unified processing of the single-cell transcriptomics atlas and hierarchical cellular taxonomy. **(a)** Hierarchical structure of transcriptome-based cellular taxonomy. Taxonomic annotation at three levels of granularity; class (n=8), subclass (n=27), and subtype (n=65). **(b)** Spatial distribution of major cell classes. **(c)** Spatial distribution of EN subclasses. **(d)** Markers defining neuronal subclasses. **(e)** Functional enrichment of cellular subclasses using Gene Ontology Biological Process (GO BP). **(f)** Enrichment of heritable traits for cellular subclasses using previous GWAS studies (scDRS). AD^11^, MS: multiple sclerosis^12^, PD^13^, Epilepsy: epilepsy focal^14^, Migraines^15^, Stroke^16^, ALS: amyotrophic lateral sclerosis^17^, SCZ^18^, BD^19^, MDD: major depressive disorder^20^, ASD: autism spectrum disorder^21^, ADHD: attention deficit hyperactivity disorder^22^, Insomnia^23^, Education: educational attainment^24^, IQ: intelligence^25^, Alcoholism^26^, OCD: obsessive compulsive disorder^27^, Tourettes: Tourette Syndrome^28^, Obesity^29^, T2D: type 2 diabetes mellitus^30^, Cholesterol: cholesterol total^31^, RArthritis: rheumatoid arthritis^32^, IBD: inflammatory bowel disease^33^, UC: ulcerative colitis^34^.

### Inter-individual variation of the human cortical transcriptome

Population-level transcriptomic variation is influenced by genetic differences among individuals, phenotypes such as age, sex, and disease status, as well as technical factors such as tissue source, dissection bias, and single-cell library preparation. We first set out to explore the determinants of the overall transcriptome variation in our sample cohort. Population-scale snRNA-seq data allowed partitioning of the gene expression variance by cellular variables (including cell type and fraction of mitochondrial and ribosomal genes), donor-level variables (including subject ID, age, sex, genetic ancestry, and diagnosis), and technical variables (including source of the tissue specimen, post-mortem interval, technical replicates, and sequencing depth) (**Fig. 3a**). Across all genes, a mean of 49.8% of the total expression variance can be attributed to variation across cell type, while inter-individual variation explains 9.8%, and unexplained residual variation accounts for 38.9%. The remaining variables explained less than 1% of the total variance on average. We prioritized several drivers of expression variation (**Fig. 3b**). Several genes with high cell type variation (including ATP8A2, GABRB3, and DOCK3) are dominated by genes differentially expressed between neuronal and non-neuronal cell types (**Fig. 3c**). As expected, genes varying across sexes were located in sex chromosomes (**Supplementary Fig. 3c**). Interestingly, genes with high variation across tissue sources were mostly mitochondrial genes (**Supplementary Fig. 3d**), possibly due to technical differences in physiological and environmental factors from dissection and handling at the respective tissue sources^35,36^. Top 3 genes with the highest variation across diagnosis (CIRBP, FGF1, and WSB1) were implicated in stress response and hypoxia. CIRBP, which also had a high variation across PMI (ranked 4th), was a gene induced in response to low temperature and hypoxia, and its expression was inversely associated with patient survival in cancer^37^. FGF1 was another gene linked to temperature stress and hypoxia^38^, and WSB1 was a neuroprotective protein and regulator of many genes associated with the cellular response to hypoxia^39^. Top variable gene in PMI, HP1BP3, was chromatin organizing protein induced in hypoxic conditions^40^. This analysis examines expression variation shared across cell types, so the low variance fraction explained by diagnosis indicates the need for cell type specific analysis.

**Fig. 3.**
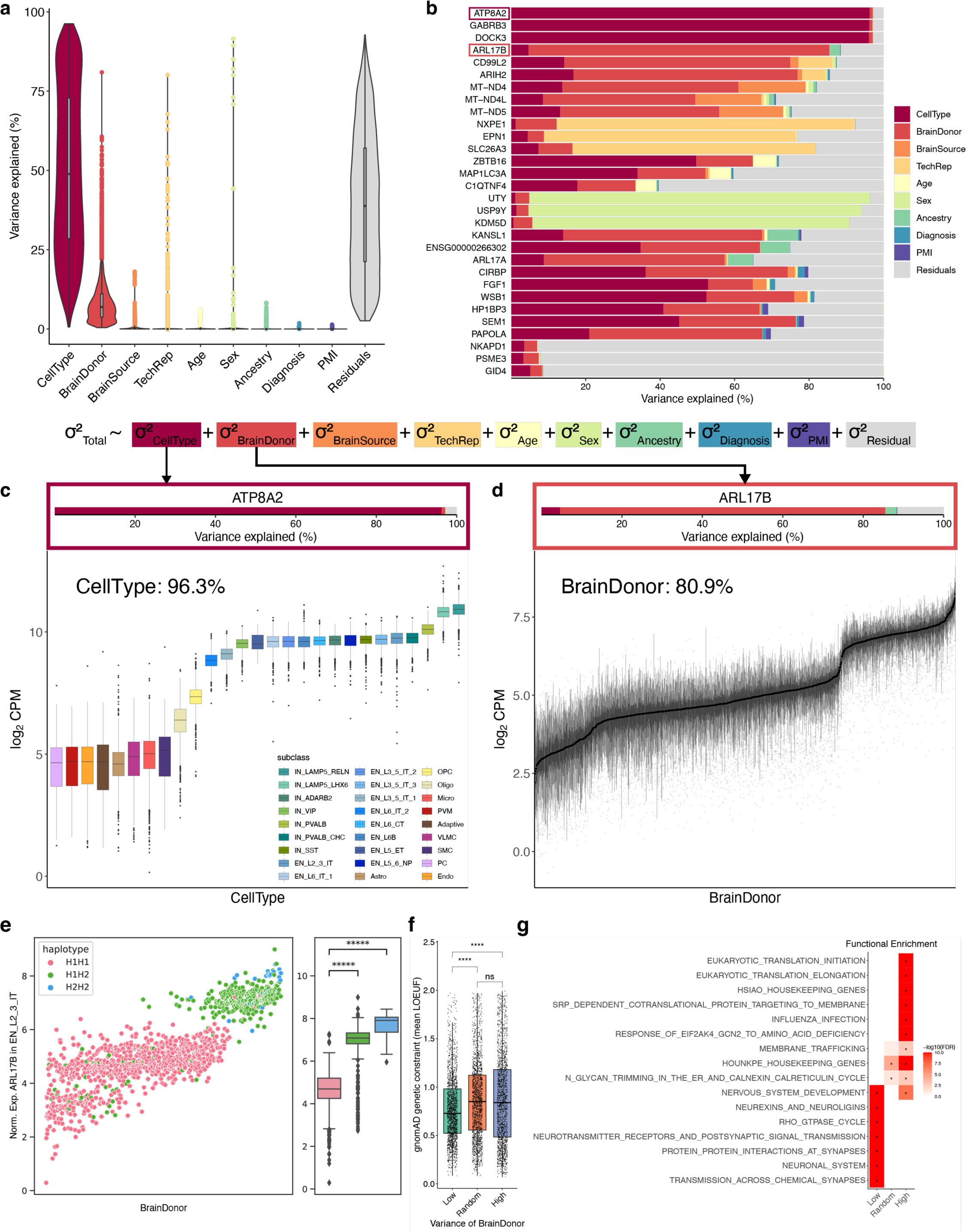
Sources of transcriptomic variation. **(a)** Variance partition of the transcriptome. **(b)** Top genes for each technical and clinical covariate category. **(c)** ATP8A2 gene expression across cell types. **(d)** ARL17B gene expression across donors. **(e)** Stratification of ARL17B expression by MAPT haplotypes. **(f)** Genetic constraints of gene groups measured by average LOEUF score (upper bound of 90% confidence interval for o/e ratio for high confidence pLoF variants; lower values indicate more constrained). “Low”, indicates the bottom 2,000 genes with the lowest brain donor variance. “High”, indicates the top 2,000 genes with the highest brain donor variance. “Random”, indicates 2,000 random genes that are in neither the “Low” nor “High” category. **(g)** Functional enrichment for genes with high or low inter-individual variation.

We observed that inter-individual differences explained 80.9% of the variation in ARL17B gene expression (**Fig. 3b,d**). Several adjacent genes, such as ARL17A and KANSL1, also have high inter-individual variation and are localized on the disease-associated MAPT locus (17q21.31) (**Fig. 3b**). ARL17B and KANSL1 are often found as a fusion transcript (KANSL1::ARL17B) and frequently undergo polymorphic translocation^41,42^, and they have been implicated in neurological disorders such as ALS, PD, and MS^43–45^. To interrogate possible genetic causes for variation in expression, we examined normalized gene expression at the donor level, stratified by MAPT haplotypes (**Fig. 3e**, **Methods**). We observed two distinct patterns of ARL17B expression that could be potentially linked to H1 and H2 MAPT haplotypes, with lower expression linked to the H1H1 genotype. We observed stratification of haplotypes by genetic ancestry, where the H2H2 genotype was almost exclusively found within EUR ancestry, consistent with previous reports^46^ (**Supplementary Fig. 3f**). We replicated a previous finding that the H1 haplotype is associated with PD susceptibility, and observed that the H1H1 genotype increases PD risk with an odds ratio (OR) of 4.125 (**Supplementary Fig. 3g**; P ≤ 0.0273), much higher than previously reported (1.42^47^ or 1.46^48^). In addition, we tested the contribution of the H1 haplotype to AD among non-ApoE4 carriers^49^ but did not find a significant association (P ≤ 0.302). The variation in inter-individual expression is inversely correlated with genetic constraints, as measured by the gnomAD Loss-of-function Observed/Expected Upper-bound Fraction (LOEUF) score (**Fig. 3f**, **Supplementary Fig. 3h**). Genes with high inter-individual variability tend to be less constrained and are often associated with the maintenance of basal cellular functions (i.e., housekeeping genes), including translation, RNA processing, metabolism, signal transduction, and structural maintenance, consistent with previous findings^50,51^ (**Fig. 3g**).

### Cross-disorder variation of cell type composition across 8 different neurodegenerative and neuropsychiatric diseases

Utilizing 318 neurotypical donors as a baseline, we systematically evaluated variation in the cellular composition of the DLPFC across eight different neurodegenerative (NDDs, including AD, DLBD, Vas, Tau, PD, and FTD) and neuropsychiatric diseases (NPDs, including SCZ and BD). Using all subclass-level cell types, we found the overall cell type composition changes were broadly stratified by NDDs and NPDs, and that they form distinct clusters (**Fig. 4a**). Notably, we observed a higher degree of similarity among AD, DLBD, and Vas. Focusing solely on neurons, we saw equal or greater correlations among the same class of diseases, underscoring the critical role of neurons in the etiology of neurological diseases. Additionally, similarities between FTD-AD (all cells) and FTD-Tau (neurons) were observed. Exploring each subclass further, we identified a notable overlap in the prevalence of neuronal and glial cell types within the same class of diseases (**Fig. 4b, Supplementary Table 3**). Specifically, we observed that NDDs were characterized by a higher abundance of non-neuronal cells, particularly vascular cell types (**Supplementary Fig. 4c**), as well as elevation of a specific IN, namely IN_LAMP5_RELN, IN_LAMP5_LHX6, and IN_ADARB2 subclasses. In contrast, NPDs were predominantly associated with an increase in neuronal cells, particularly deep layer ENs in L5-6. To further identify specific subtypes responsible for driving the compositional changes in vascular cell types, we used subtype-level annotation to analyze compositional variation in 8 NDDs and NPDs (**Fig. 4c, Supplementary Fig. 4d**). From this, we identified dominant subtypes of each subclass that further differentiated NDDs and NPDs. For example, vascular leptomeningeal cells (VLMCs) are barrier-forming fibroblasts of the brain^52^, and they are transcriptionally segregated into three subtypes; two meningeal VLMCs (VLMC_DCDC2 and VLMC_SLC4A4) and one perivascular VLMC (VLMC_ABCA6). Our subtype-level analysis indicates a polarized response of meningeal VLMC_DCDC2 where their increased proportions are specifically associated with most NDDs.

**Fig. 4.**
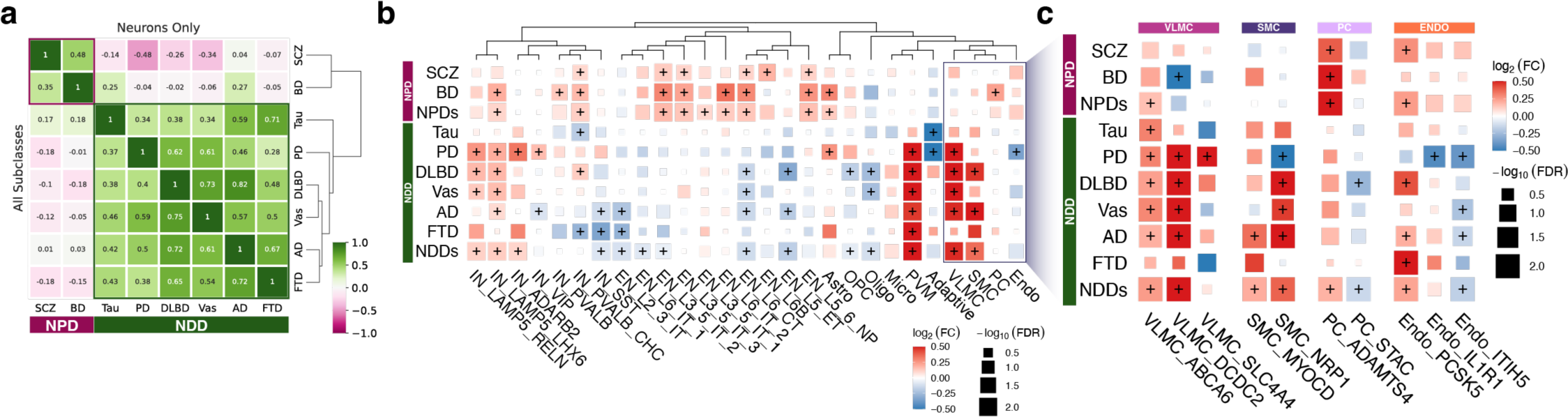
Cross-disorder variation of cell type composition comparing 8 different NDDs (AD, DLBD, Vas, Tau, PD, and FTD) and NPDs (SCZ and BD) against common neurotypical controls. **(a)** Correlation of cell type composition using all cell types (bottom-left triangle) or limited to neuronal cell types (upper-right triangle). **(b)** Variation in cell type composition for each subclass in 8 different diseases. NDDs and NPDs indicate meta-analysis using broad disease categories. **(c)** Variation of subtype-level composition in the vascular cell class. Color intensity indicates effect size and dot size reflects the statistical significance of correlations.

### Cross-disorder variation of gene expression and genetic concordance

Cross-disorder gene expression analysis has the potential to identify shared biological pathways and mechanisms, improve diagnostic accuracy, and help develop targeted treatments. To assess the extent of sharing between NDDs and NPDs, we performed a comprehensive analysis of differentially expressed genes (DEGs) by decomposing the total disease signatures into shared and distinct components. Using Dreamlet followed by Mashr^53^, we performed composite tests to evaluate the specificity of disease effect (**Methods**), leading to the identification of shared disease signatures that are invariant across cell subclasses (**Fig. 5a, Supplementary Figs. 5a,b**). Genes sharing cross-disorder signatures encompassed crucial transcriptional processes, such as mRNA splicing and processing, and protein localization to mitochondria. The observed cross-disorder signatures, affecting genes, which are critical for the proper functioning of cellular processes, align with the omnigenic model^54^, and further support the pleiotropy of those genes influencing multiple disorders, both genetically and transcriptionally. After discounting the shared cross-disorder signatures from the overall DEG expression profiles, we used the residual effects to quantify the pair-wise transcriptomic similarity between traits (**Fig. 5b, Supplementary Table 4**). Similarities between pairs of NDDs or NPDs were greater compared to the NDD-NPD contrast, with AD, DLBD, Vas, and PD being the most similar. Meta-analysis using disease-specific effects of these four transcriptomically similar traits (AD, DLBD, Vas, and PD) implicated neuronal development and synaptic signaling pathways involving interneurons (IN_LAMP5_RELN, IN_ADARB2, and IN_PVALB) as well as vasculature development from the VLMC subclass (**Fig. 5c, Supplementary Fig. 5c**).

**Fig. 5.**
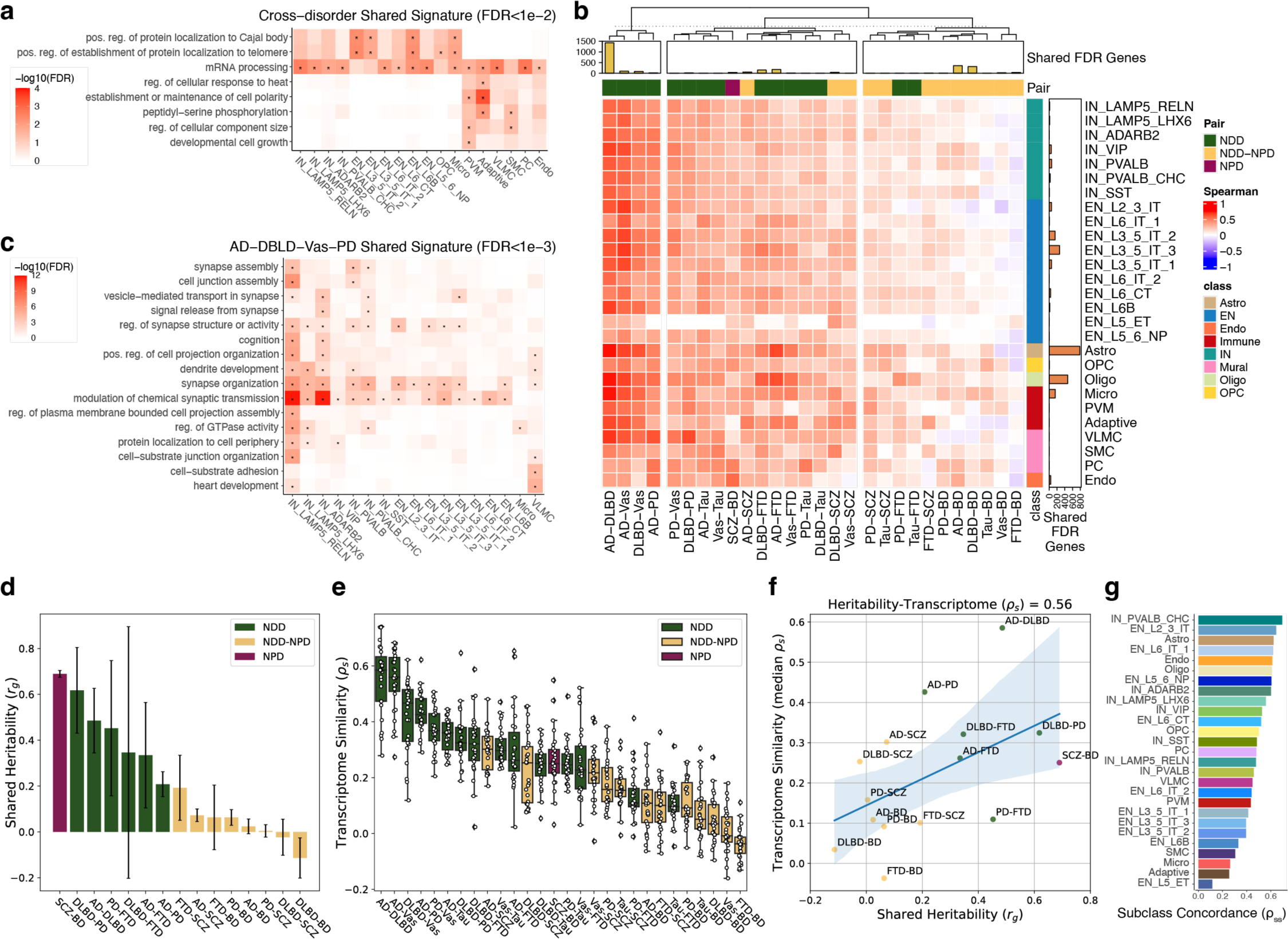
Cross-disorder variation of gene expression. **(a)** Pathways implicated by shared gene expression changes across 8 disorders. Hypergeometric test with FDR ≤ 0.01 shown. **(b)** Transcriptome similarities between disease pairs. Correlation is measured using Spearman correlation. Cross-disease shared genes are discounted from the comparison. **(c)** Pathways implicated by shared signatures from AD, DLBD, Vas, and PD. Hypergeometric test with FDR ≤ 0.001 shown. **(d)** Shared heritability estimates for disease pairs. LD Score Regression (LDSC). **(e)** Transcriptome similarity measured by Spearman correlation. **(f)** Correlation between shared heritability and median of transcriptome similarity. **(g)** Correlation between shared heritability and transcriptome similarity per cell subclass.

Comparison of pairwise trait co-heritability against pairwise transcriptome similarity can identify shared genetic influences and underlying biological mechanisms, leading to better insights into disease etiology and potential therapeutic targets. We first used GWAS summary statistics to estimate the shared heritability (r_*g*_) among NDDs and NPDs, revealing disease pairs (i.e., SCZ-BD, DLBD-PD) with a high degree of heritability overlap (**Fig. 5d**). This analysis differentiates between NDDs and NPDs, revealing lower shared heritability between NDDs and NPDs. We then quantified the pairwise transcriptome similarity (ρ_s_) across NDDs and NPDs (**Fig. 5e**). Our findings revealed varying degrees of transcriptomic overlap, with some exhibiting high similarity, particularly in disease pairs such as AD and DLBD. We calculated the pair-wise trait heritability-transcriptome concordance (*r_g_-ρ_s_*) by comparing the shared heritability against the average transcriptome similarity across cell types. We observed a positive correlation between genetic and transcriptomic similarities (Spearman’s *ρ*=0.56), indicating that diseases with higher shared genetic risk also tend to have more similar gene expression profiles (**Fig. 5f**). We extended the *r_g_-ρ_s_* comparison by considering the transcriptional concordance for each cell type (**Fig. 5g, Supplementary Fig. 5d**). Among all cell types, the transcriptional concordance of Chandelier cells (IN_PVALB_CHC) had the highest similarity with the pairwise trait heritability. In summary, our approach allowed us to dissect disease signatures into shared and distinct components, revealing significant overlap in gene expression and genetic risk across NDDs and NPDs. These findings bolster our understanding of common and unique disease mechanisms, paving the way for novel therapeutic strategies targeting shared or distinct pathways.

### Transcriptomic variation with AD pathology

Next, we focused on characterizing the transcriptomic variation in AD using case-control comparison, and analysis of different phenotypes that capture disease severity, including plaque density using CERAD scores, neurofibrillary tangle progression using Braak stage, and level of cognitive impairment. The analysis of variation in cell type composition reveals distinct patterns associated with AD pathology compared to normal aging (**Fig. 6a, Supplementary Fig. 6a, Supplementary Table 5**). Notably, changes unique to AD pathology, such as the observed increase in Micro and IN_LAMP5_LHX6 with higher CERAD scores, indicate a potential association between this neuronal subtype and Aβ pathology. Additionally, the loss of EN_L2_3_IT in AD (based on case-control and CERAD comparisons), which is not evident in normal aging, suggests a specific vulnerability for this neuronal subtype to AD. Placing subclasses based on compositional shifts of normal aging and AD (**Fig. 6b**), we identified one of the vascular cell types, SMC, was having the opposite effect, suggesting an AD-specific vulnerability in SMC. Likewise, changes in most ENs were discordant suggesting the mechanisms that lead to neuronal loss are AD-specific. Taken together, our findings highlight the cell type-specificity of vulnerability associated with AD.

**Fig. 6.**
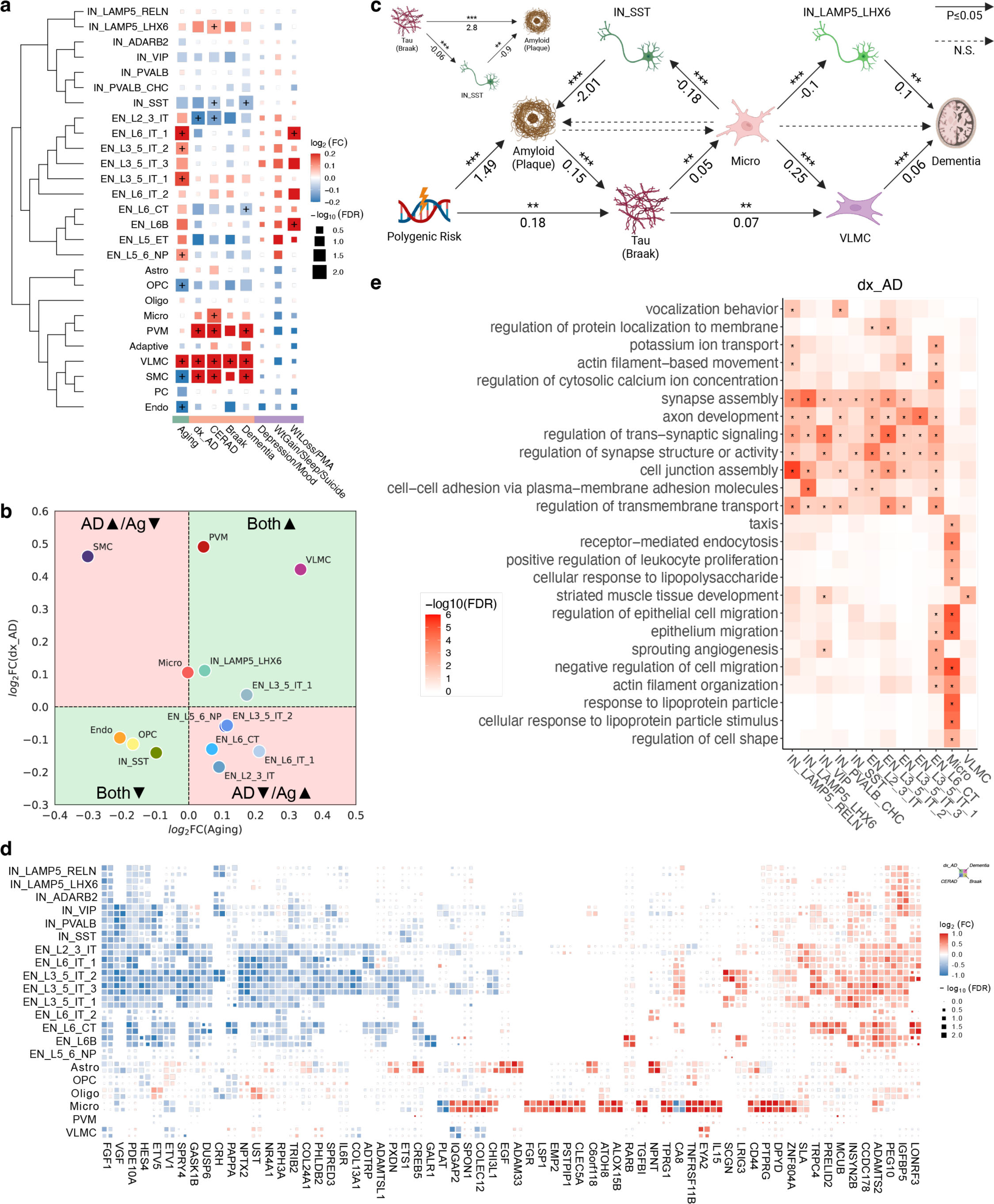
Transcriptome changes across AD neuropathology. **(a)** Compositional variation analysis using normal aging, different measures of AD pathology (binary AD diagnosis (dx_AD), CERAD score, Braak staging, and ordinal dementia scale), and 3 categories of NPS within AD. **(b)** Comparison of compositional changes between aging and AD. Green indicates changes are concordant. Red indicates changes are discordant. Only subclasses with at least one FDR significant contrast shown. **(c)** Causal mediation analysis using PRS, mean plaque, Braak staging, and dementia scale. CLR-transformed subclass fractions were used for modeling. Statistical significances are shown above the arrow (P < 0.001 ***, < 0.01 **, and < 0.05 *), and numbers below indicate coefficients. Mediation effects of SST interneurons on Aβ plaque accumulation shown separately. **(d)** DEGs in AD phenotypes. Meta-analysis between brain banks. Top genes with FDR < 0.01 and effect size ≥ 0.35. **(e)** Functional enrichment analysis of DEGs by subclass using Gene Ontology Biological Process. Hypergeometric test with FDR ≤ 0.01 shown. GO terms were reduced using rrvgo.

To better understand transcriptomic variation under AD pathology and to identify subclass most affected, we also analyzed the changes in AD cases due to co-occurring conditions. It is estimated that more than 80% of AD patients will exhibit at least one NPS over the course of their illness that significantly impacts their clinical outcomes^5,55^ which suggests at least some shared molecular mechanisms between serious mental illness and AD. To evaluate cell type associations with the prevalence of NPS, we applied compositional variation analysis to three categories of NPS based on co-occurrence estimates. Notably, using age-matched groups of AD patients with or without NPS (**Supplementary Fig. 6b**), we found AD patients experiencing weight loss and PMA have an increased ratio of EN cell types, especially in deep layer neurons in L6 (**Fig. 6a, Supplementary Fig. 6a**). Our results support previous findings^56^ that the deeper PFC layers (L5-6) are involved with certain types of NPS.

Upon identification of vulnerable cell subclasses in AD, we questioned their roles in AD pathology whether their changes are damaging, protective, causal, or derived. As demonstrated in previous studies^57^, we employed mediation analysis to decipher the causal relationships between various cascades of events leading to disease onset and progression of AD pathology. We performed the analysis on the base hypothesis that polygenic risk (AD PRS; **Methods**) contributes to plaque accumulation (mean density of neuritic plaques), which in turn affects tau progression (Braak), leading to varying degrees of dementia. We tested 14 subclasses that were significantly altered in any of the contrasts and observed significant average causal mediation effects (ACME, P < 0.05) involving Microglia, VLMC, IN_SST, and IN_LAMP5_LHX6 (**Fig. 6c**). Initially, we showed PRS affects both plaque and tau pathology, but tau is also significantly affected by plaque accumulation (ACME = 0.0903, P < 2e-16). Then, tau progression results in more VLMC cells mediated by an increase in Microglia (ACME = 0.00477, P = 0.005). Conversely, an increase in microglia leads to a decrease IN_LAMP5_LHX6. Such changes in the levels of both VLMC (ACME = 0.00515, P = 0.0002) and IN_LAMP5_LHX6 (ACME = −0.00269, P = 0.031) appear to exacerbate dementia. Furthermore, plaque accumulation is mitigated by IN_SST cells (ACME = 0.1475, P < 2e-16). Increased microglia lowers IN_SST (coef = −0.18, P = 5.04e-07), as does increased tau (coef = −0.06, P = 1.82e-04), contributing to more plaque accumulation (coef = −2.01, P = 7.47e-06).

To determine how gene programs change in response to increasing severity of AD pathology, we characterized the DEGs in 27 subclasses (**Fig. 6d, Supplementary Fig. 6d, Supplementary Table 6**). DEGs in general have high concordance across different AD pathology variables, consistent with previous reports^1^. Overall, DEGs in AD (FDR < 0.05) can be summarized as up-regulation of genes in vascular cell classes (Mural and Endo) while down-regulation of genes in neurons (**Supplementary Fig. 6c**). We discovered genes differentially expressed in microglia were not observed in other subclasses, and these genes generally exhibited higher effect sizes. These include previously characterized up-regulated genes in Microglia, including DPYD, IL15, and PTPRG^53,58,59^. Further characterizing microglia-associated gene signatures, we found they are specifically enriched with pathways involved in negative regulation of cell motility and migration as well as response to lipoprotein particle (**Fig. 6e, Supplementary Fig. 6e**). Gene expression changes in neurons were largely affecting synaptic functions including synapse assembly, development, signaling, and membrane transport. Lastly, genes affecting VLMCs were implicated in muscle tissue development.

### Nonlinear dynamics of the AD pathological trajectory

To understand AD dementia mediated by tau proteinopathy, we modeled the transcriptome using variational autoencoder (VAE)-based latent manifold mapping (**Fig. 7a, Methods**). We inferred two independent cell-type-specific disease trajectories from semi-quantitative measures of AD progression (tau proteinopathy and severity of cognitive decline) and explored the dynamics of two paths leading to the onset and early stage of AD pathogenesis. To decorrelate Braak and dementia model predictions, we equally sampled all combinations of Braak stage and dementia status during model training (**Supplementary Fig. 7**). The accuracy of the Braak and dementia model predictions were significantly above chance for all eight cell classes (P < 1e-4, bootstrap) (**Fig. 7b, top**). We built a disease trajectory using the predicted Braak staging as a pseudo-temporal axis (**Supplementary Figs. 8a,b**) and calculated a dementia resilience score that measured how gene expression is correlated with predicted dementia, conditioned on the predicted Braak staging (**Methods**). Specifically, a gene is considered protective (i.e., resilient against dementia), if increased expression is associated with a decrease in predicted dementia, and vice versa (damaging if increased expression is associated with an increase in predicted dementia). Many past studies have suggested that gene expression can evolve nonlinearly with disease progression^60^. Thus, we measured the degree of nonlinearity for each gene trajectory (**Supplementary Fig. 8c**) and revealed that those within the immune and neuronal cell class were the most nonlinear and linear, respectively (both Ps < 1e-4, bootstrap) (**Fig. 7b, bottom**).

**Fig. 7.**
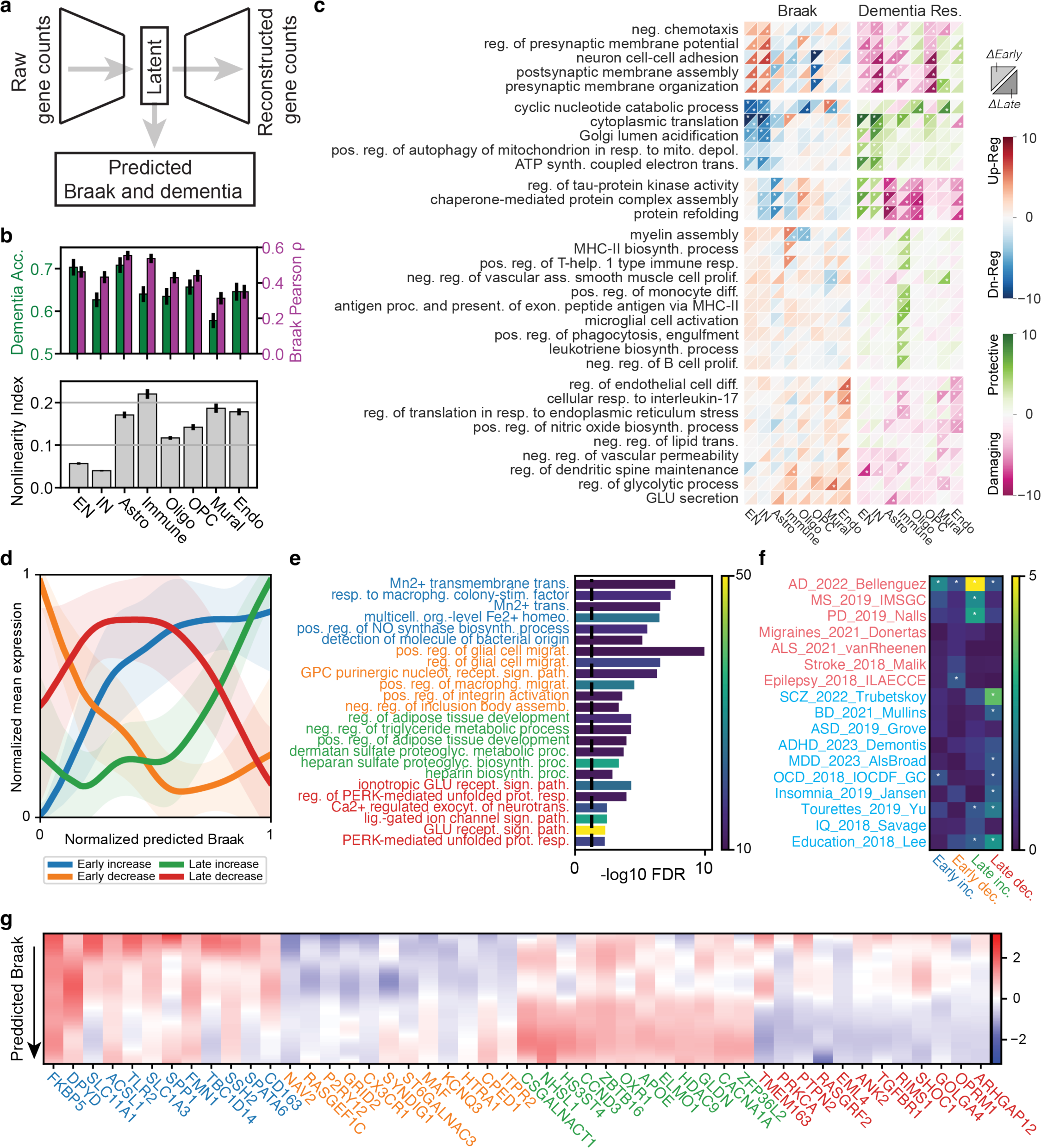
Modeling of AD using disease trajectory analysis. **(a)** Overview of the trajectory inference model. **(b)** Accuracy of the model (above) and nonlinearity index by class (below). **(c)** Pathway enrichment by early (upper left triangles) and late (lower right triangles) phases of the AD trajectory, as predicted by Braak and dementia resilience. A gene is considered protective if increased expression is associated with a decrease in predicted dementia, and vice versa. Hue indicates the z-score (clipped between −10 and 10), and stars indicate FDR < 0.05. **(d)** Four representative disease trajectory modules for immune cell class. The mean normalized expression of the 250 genes with greatest early increasing (blue curve), early decreasing (orange), late increasing (green) and late decreasing (red) slopes based on Braak trajectories. **(e)** Pathway enrichment of disease trajectory modules. Text colored by the four trajectory modules. Hue indicates the number of genes in the pathway. **(f)** Enrichment of heritability estimates (MAGMA) for each disease trajectory module. Text color indicates neurological traits (red) and psychiatric traits (blue). Hue indicates negative log10(FDR) (clipped at 5), asterisk indicates FDR < 0.05. **(g)** Top 12 genes for each trajectory module. Hue indicates the rate of normalized expression change (slope) based on the predicted Braak staging. For visualization, we only show each gene once even if it appears in more than one module.

To further characterize the nonlinear evolution of gene expression, we defined an “early” and “late” disease stage (**Supplementary Fig. 8c**) and determined the top 32 GO BP pathways that best summarize the biological changes associated with early and late stages of tau proteinopathy and dementia resilience (**Fig. 7c**, Supplementary Figs. 9-11**, Supplementary Table 7)**. A semantic clustering of the pathways identified five functional clusters. The first cluster was primarily related to synaptic function, which were characterized by down-regulation in OPCs in the early stages of AD, and then up-regulation in neurons in later stages. For both cell types, increased expression was associated with diminished resilience (i.e., damaging). The late increase of genes implicated in these pathways could be linked to compensatory mechanisms following synaptic loss^61^. The second cluster was related to cell metabolism pathways implicated in protein translation^2,62^, mitochondrial function^2,63^, and acidification^64^, which showed strong downregulation in neurons with increasing Braak, consistent with past studies^65^. Decreased expression in these metabolic pathways implied damaging association with decreased resilience (increase in metabolism was protective). The third cluster was related to cell stress, including pathways related to chaperone-mediated protein assembly, folding, and tau-kinase activity. In non-neuronal cells, increased cell stress was strongly associated with cognitive decline in both the early and late stages. The fourth cluster was related to immune response and inflammation. In immune cells, early increase in pathways such as microglial activation, phagocytosis, and B cell proliferation was observed, while late increase was involved with pathways implicated in adaptive immune responses such as antigen presentation and T-cell response, all of which were associated with dementia resilience. Lastly, vascular cells were implicated in damaging changes such as reduced lipid transport and vascular permeability, response to interleukin-17 (IL-17), glycolysis, endothelial cell differentiation, and nitric oxide synthesis.

As previous studies have highlighted the importance of the immune response in AD^58,66–72^, we examined this cell class in more detail (**Figs. 7d-g**); We also extended the analysis to EN, IN and OPC cells classes (**Supplementary Figs. 12-15**). To help visualize the non-linear dynamics of the immune response to AD, we categorized gene expression trajectories into four modules based on their response to increasing tau proteinopathy: early increasing, early decreasing, late increasing, and late decreasing (**Fig. 7d**). Gene enrichment of these four trajectory modules showed that pathways involved in macrophage colony stimulation and metal transport were all upregulated in the early stage (**Fig. 7e**), whereas migration, purinergic signaling, and negative regulation of inclusion body assembly were downregulated. In the later stage, lipid-related pathways such as adipose tissue development, and triglyceride metabolism increased, while synapse-related pathways were downregulated. Gene-set enrichment analysis using GWAS summary statistics for the top 250 genes in each trajectory module revealed that late increasing genes were the most strongly associated with AD (FDR ≤ 4.1e-6), although the other three modules were also significantly associated (FDR early increase ≤ 0.005, early decrease ≤ 0.048, late decrease ≤ 0.040 (**Fig. 7f, Supplementary Table 8, Supplementary Fig. 12b**). Interestingly, the late increasing trajectory module was also significantly correlated with other NDDs, including MS and PD. We further examined the genes associated with trajectory modules (**Fig. 7g**, list of top genes for all cell classes are found in **Supplementary Table 9**). Markers for homeostatic microglia^73^, such as CX3CR1, NAV2 and P2RY12 were among the top 10 early decreasing genes with increasing proteinopathy (FRMD4A ranked 26th out 17,265 coding genes, **Supplementary Table 9**). Conversely, some of the early increasing genes, such as ACSL1, DPYD, and CD163 were recently implicated in a pathogenic lipid-droplet accumulation phenotype in individuals with AD who carry the APOE4/4 genotype (another implicated gene, NAMPT, ranked 54th. **Supplementary Table 9**)^70^. These genes, along with the later upregulation in lipid-related pathways such as adipose tissue development and triglyceride metabolism, lend support to the hypothesis that microglia develop a lipid-droplet accumulating state that potentially exacerbates disease progression^70–72^.

## Discussion

We report a comprehensive disease atlas of the human DLPFC using 1,494 donors affected with various complex neurological and psychiatric conditions. Our single-nucleus transcriptomic analyses provide novel insights about cellular heterogeneity and variability in the human brain, including the observation that about 10% of total transcriptomic variation can be attributed to inter-individual differences. Intriguingly, we find genes with higher inter-individual variability and lower genetic constraints that were implicated in certain housekeeping roles^51^. This is in contrast to the idea that, due to their essential nature, housekeeping genes are more likely to be conserved^74,75^. They are among the most universal genes in the cell, and cells adapt their protein synthesis machinery in response to physiological needs^76^. This flexibility is crucial for processes such as differentiation, proliferation, and response to stress. A higher degree of regulatory flexibility suggests a tolerance for variation within certain constraints.

We used the disease atlas to characterize cellular and disease-specific responses to pathologic conditions. Our findings reveal shared and distinct cellular composition profiles among NDDs and NPDs, furthering our understanding of their underlying pathophysiological mechanisms. Disease signatures shared across NDDs and NPDs are enriched with genes critical for the proper functioning of cellular processes, such as RNA splicing. This observation aligns with the omnigenic model^54^, where most heritability in complex traits can be explained by effects on peripheral genes that often play indirect, subtle, and cumulative roles in disease. Thus, discounting cross-disease effects could facilitate identification of core disease-relevant functional genes. Using this approach, we found that disease pairs with higher genetic risk overlap tend to have greater transcriptomic concordance, in a cell-type-dependent manner, suggesting that genetic factors contributing to disease susceptibility can also influence transcriptomic alterations in similar ways^3^.

In addition, we revealed that the brain vascular system is intricately linked to immune dysfunction in NDDs. In general, we saw a relative increase in vascular cell types in most NDDs and demonstrate that, in a seemingly protective role, levels of VLMCs rise in individuals who experience exacerbated cognitive impairment in AD. It has previously been shown that the meningeal lymphatic system plays multiple roles in the brain, including waste removal^77–79^ and the adaptive immune response^52,80^. Given the role of VLMCs in these processes^81–84^, they warrant further investigation in the context of NDDs.

Our analysis of the pathological trajectory of AD aligns with various proposed hypotheses^65,85–89^, particularly for pathways affected in the earliest stages of the disease. Neuronal, immune, and vascular cells exhibit distinct vulnerabilities, with shared alterations across correlated pathways. In neurons, we observed a decrease in metabolic functions (cyclic nucleotide catabolic process, cytoplasmic translation, and ATP synthesis) that have been closely linked to synaptic dysfunction and cognitive decline^65^. The immune response was generally protective against cognitive decline: an early innate immune activation^85^ followed by an adaptive immune response^67–69^ was associated with dementia resilience. The one exception was the response to IL-17, which was damaging in the early stages of AD for the Immune, Mural, and Endo classes. IL-17 has been associated with cognitive decline^90,91^ and disruption of the blood-brain barrier^92^, with anti-IL-17 treatment restoring cognitive function in mice^90,91^. In addition, chaperones have been closely associated with a number of disease related processes, including tau misfolding and aggregation, and neurotoxicity^86–89,93,94^, however, their precise roles in AD pathogenesis remains unclear. Our results suggest that chaperones can play opposing roles; protective for neurons but damaging for glia and immune cells. Lastly, vascular cells are associated with negative regulation of vascular permeability and lipid transport, which are detrimental to the infiltration of perivascular immune cells^95^ and the clearance of protein aggregates. Taken together, these insights deepen our understanding of AD pathogenesis and implicate cellular responses that warrant further investigation. Overall, the PsychAD single-cell disease atlas serves as a unique and foundational resource to further our understanding of population-level disease-associated transcriptomic variation in the human brain.

## Methods

### Collection and harmonization of clinical, pathological, and demographic metadata

Brain tissue specimens were sourced from two brain banks: the Mount Sinai NIH Neurobiobank (MSSM; 1,042 samples) and the NIMH-IRP Human Brain Collection Core (HBCC; 300 samples). Additionally, samples were obtained from five prospective cohort studies conducted at the Rush Alzheimer’s Disease Center (RADC; 152 samples)^96,97^. As such, the available clinical data varied as a function of source (**Supplementary Fig. 4a**). We used the following scheme to harmonize available clinical, pathological, and demographic metadata: the CERAD scoring scheme for neuritic plaque density^98^ was harmonized for consistency across multiple brain banks, where the scores range from 1 to 4, with increasing CERAD number corresponding to an increase in AD burden; 1=no neuritic plaque (normal brain), 2=sparse (possible AD), 3=moderate (probable AD), 4=frequent (definite AD). Samples from RADC used consensus summary diagnosis of no cognitive impairment (NCI), mild cognitive impairment (MCI), and dementia and its principal cause, Alzheimer’s dementia^99–101^. MSSM samples used clinical dementia rating (CDR), which was based on a scale of 0-5; 0=no dementia, 0.5=questionable dementia (very mild), 1=mild dementia, 2=moderate dementia, 3=severe dementia, 4=profound dementia, 5=terminal dementia. After consulting with clinicians, we created a harmonized ordinal variable where dementia is categorized into three levels of cognitive decline, independent of AD diagnosis; 0=no cognitive impairment, 0.5=MCI (mild cognitive impairment), and 1-5=dementia. In addition to AD phenotype, we collected comprehensive demographic (age, sex, and genetic ancestry) and technical variables (tissue source, technician, sample batch, postmortem interval (PMI; measured in minutes), ApoE genotype) to describe each cohort (**Supplementary Table 1**). We briefly describe the process for assigning genetic ancestry^102^. In particular, we leveraged quadratic discriminant analysis (QDA) to infer genetic ancestry by training our model using data from the 1000 Genomes Project. We utilized 10-fold stratified cross validation to optimize the regularization parameter within QDA^103^ as well as forward selection to identify the optimal number of principal components for genetic ancestry assignments. For samples without genotypic data we utilized race/ethnicity as a proxy for inferring genetic ancestry. We emphasize that while genetic ancestry is a distinct concept from the social constructs of race and ethnicity^104^, we leveraged the correlated race/ethnicity variables as proxies to retain those samples in the analyses. Values for superpopulations included: EAS, SAS, AFR, AMR, EUR, and EAS_SAS, where the category “EAS_SAS” was assigned for samples with unavailable genotypes with an “Asian” value for race/ethnicity, which can potentially correspond to both EAS and SAS.

### Clinical diagnosis of AD

For analysis comparing donors with AD cases and neurotypical controls, a binary clinical diagnosis variable for AD, dx_AD, was defined, as follows: Individuals with CERAD 2, 3, or 4, Braak ≥ 3, and CDR ≥ 1 for MSSM or Alzheimer’s dementia for RADC were classified as AD cases. Controls were defined as individuals in *controls_neuropathological_clinical* category where CERAD={1}, Braak={0,1,2,3}, and secondary diagnosis (including dementia) is not allowed except for MCI.

### Measuring AD neuropathology

For analysis comparing donors with pathologic AD, the following variables were used to measure the severity of AD neuropathology: **CERAD score**^98^. A quantitative measure of Aβ plaque density where 1 is normal, 2 is possible AD, 3 is probable AD, and 4 is definite AD^99^. **Braak AD-staging score** measuring progression of neurofibrillary tangle neuropathology (Braak & Braak-score, or BBScore). A quantitative measure of the regional patterns of neurofibrillary tangle (NFT) density across the brain, where 0 is normal and asymptomatic, 1-2 indicate initial stages where NFT begins to appear in the locus coeruleus and the transentorhinal region, 3-4 indicate progression to limbic regions, such as the hippocampus and amygdala, and 5-6 indicate NFT are widespread, affecting multiple cortical regions^105–107^.

### Measuring cognitive impairment

For analysis comparing donors with AD-related dementia, the following variable was used to measure the severity of cognitive impairment: **Clinical assessment of dementia.** A harmonized variable of cognitive status based on the CDR scale for MSSM or NCI, MCI, and Alzheimer’s dementia for RADC. We used the three-level ordinal categories of clinical dementia to measure the severity of dementia, in which 0 indicates no dementia, 0.5 indicates minor cognitive impairment, and 1.0 indicates definite clinical dementia.

### Definition of cross-disorder contrasts

For cross-disorder contrasts, we limited the analysis to any individual with Age ≥ 17. Neurotypical controls are defined as any individual CERAD={1}, Braak={0,1,2,3}, and secondary diagnosis is not allowed. AD is any individual with CERAD={2,3,4}, Braak={3,4,5,6}, clinically diagnosed as dementia, and secondary diagnosis not allowed. SCZ is any individual with SCZ diagnosis (*SCZ* | *Schizoaffective_bipolar* | *Schizoaffective_depressive*) and secondary diagnosis not allowed, except for metabolic and eating disorders. DLBD is any individual with DLBD diagnosis (*DLBD*) and secondary diagnosis can be only AD. Vascular is any individual with Vascular diagnosis (*Vascular*) and secondary diagnosis can be only AD. BD is any individual with BD diagnosis (*BD_unspecific* | *BD_I* | *BD_II* | *Schizoaffective_bipolar*) and secondary diagnosis not allowed except for metabolic and eating disorders. Tauopathy is any individual with CERAD={1}, Braak={4,5,6} and secondary diagnosis allowed. PD is any individual with PD diagnosis (*PD* | *PD_uncertain_plus_encephalitic*) and secondary diagnosis can be only AD. FTD is any individual with FTD diagnosis (*FTD*) and secondary diagnosis can be only AD. All disease contrasts used in the study can be found in **Supplementary Table 1**.

### Isolation and fluorescence-activated nuclear sorting (FANS) of nuclei from frozen brain specimens with hashing

All buffers were supplemented with RNAse inhibitors (Takara). 25mg of frozen postmortem human brain tissue was homogenized in cold lysis buffer (0.32M Sucrose, 5 mM CaCl_2_, 3 mM Magnesium acetate, 0.1 mM, EDTA, 10 mM Tris-HCl, pH8, 1 mM DTT, 0.1% Triton X-100) and filtered through a 40 µm cell strainer. The flow-through was underlaid with sucrose solution (1.8 M Sucrose, 3 mM Magnesium acetate, 1 mM DTT, 10 mM Tris-HCl, pH8) and centrifuged at 107,000 xg for 1 hour at 4°C. Pellets were resuspended in PBS supplemented with 0.5% bovine serum albumin (BSA). 6 samples were processed in parallel. Up to 2M nuclei from each sample were pelleted at 500 xg for 5 minutes at 4°C. Nuclei were re-suspended in 100 µl staining buffer (2% BSA, 0.02% Tween-20 in PBS) and incubated with 1 µg of a uniuqe TotalSeq-A nuclear hashing antibody (Biolegend) for 30 min at 4°C. Prior to FANS, volumes were brought up to 250 µl with PBS and 7-Aminoactinomycin D (7-AAD) (Invitrogen) added according to the manufacturer’s instructions. 7-AAD positive nuclei were sorted into tubes pre-coated with 5% BSA using a FACSAria flow cytometer (BD Biosciences).

### snRNA-seq and hashing library preparation

Following FANS, nuclei were subjected to 2 washes in 200 µl staining buffer, after which they were re-suspended in 15 µl PBS and quantified (Countess II, Life Technologies). Concentrations were normalized and equal amounts of differentially hash-tagged nuclei were pooled. Using 10x Genomics single cell 3’ v3.1 reagents (10x Genomics), 60,000 (10,000 per donor) nuclei were run in each of x2 10x Genomics lanes to create a technical replicate. At the cDNA amplification step (step 2.2) during library preparation, 1 µl of 2 µm HTO cDNA PCR “additive” primer was added^108^. After cDNA amplification, supernatant from 0.6x SPRI selection was retained for HTO library generation. cDNA libraries were prepared according to the 10x Genomics protocol. HTO libraries were prepared as previously described^108^. cDNA and HTO libraries were sequenced at NYGC using the Novaseq platform (Illumina).

### Processing of snRNA-seq data

**Alignment**. Paired-end snRNA-seq library reads were aligned to the hg38 reference genome using STAR solo^109,110^ and sample pools were demultiplexed using genotype matching via vireoSNP^111^. After per-library count matrices were generated, the downstream processing was performed using Pegasus v1.7.0^112^ and scanpy v1.9.1^113^. **QC**. We applied rigorous three-step QC to remove ambient RNA and retain high quality nuclei for subsequent downstream analysis. First, QC was applied at the cell level. Poor-quality nuclei were detected by thresholding based on UMI counts, gene counts, and mitochondrial content. We also checked for possible contamination from ambient RNA, fraction of reads mapped to non-mRNAs, like rRNA, sRNA, and pseudogenes, as well as known confounding features such as lncRNA MALAT1. Second, QC was applied at the feature level by removing features that were not robustly expressed by at least 0.05% of the nuclei. Lastly, QC was applied at the donor level by removing donors with very low nuclei counts, which can introduce noise to downstream analyses. We also removed donors with low genotype concordance. Further filtering was carried out by removing doublets using the Scrublet method^114^. **Batch correction**. We assessed the correlation between all pairs of technical variables using Canonical Correlation Analysis and used the Harmony method^115^ to regress out unwanted variables such as the effect of brain tissue source.

### Defining cellular taxonomy using iterative clustering

Cellular taxonomy was defined using a divide-and-conquer strategy. From the full dataset containing over 6 million nuclei, 8 major cell classes were defined using the following steps. We selected 6,000 highly variable genes (HVGs) from mean and dispersions trends^116^ using the default parameters (min_mean=0.0125, max_mean=3, min_disp=0.5) and brain sources as batch variable after manually excluding sex and mitochondrial chromosomes and MT. We used the k-nearest-neighbor (kNN) graph calculated on the basis of harmony-corrected PCA embedding space to cluster nuclei of the same cell type using Leiden^117^ clustering algorithms. We used UMAP^118^ to visualize the resulting clusters. From the class-level clusters, we subsetted the data by each class. Re-calculating HVGs among cells in the same class allowed us to re-focus on a feature space that is more relevant for the same class of cells. We then calculated kNN graph on the basis of the harmony-corrected PCA of the selected HVGs. Leiden-clustering was used to annotate subclass-level annotations. We iterated to the second level of taxonomy yielding 67 subtypes of human brain cells.

### Spatial validation of cellular taxonomy

#### Xenium *in situ* panel selection and custom panel design

Xenium Human Brain Gene Expression Panel (1000599, 10x Genomics) and a custom panel of 100 genes (**Supplementary Table 10**) were selected for the Xenium experiment. The 100 gene custom panel consisted mainly of subclass markers selected based on specificity and gene expression level. The custom gene list was sent to 10X genomics and the probe design was performed using their in-house pipeline.

#### Tissue preparation

Fresh frozen tissue specimens of DLPFC were dissected into small blocks on ice. Tissue blocks were snap frozen by submerging in an isopentane (320404-1L, Sigma-Aldrich) bath chilled with dry ice and stored in −80 °C. Before cryosectioning, tissue blocks were allowed to equilibrate to the cryostat (HM505, Microm) chamber temperature, and were mounted with OCT (Tissue-Tek® O.C.T. Compound, 4583, Sakura Finetek USA). After trimming, good quality 10 µm sections were flattened on the cryostat stage and placed on pre-equilibrated Xenium slides (Xenium Slides & Sample Prep Reagents, 1000460, 10x Genomics). 2-3 sections were placed on each slide. Sections were further adhered by placing a finger on the backside of the slide for a few seconds and were then refrozen in the cryostat chamber. Slides were sealed in 50 ml tubes and stored at −80 °C until Xenium sample preparation.

#### Sample preparation

Xenium sample preparation was performed according to the manufacturer’s protocol; “*Xenium In Situ for Fresh Frozen Tissues – Fixation & Permeabilization, CG000581, Rev C*” and *“Xenium In Situ Gene Expression - Probe Hybridization, Ligation & Amplification, User Guide, CG000582, Rev C’’*. Briefly, fresh frozen sections mounted on Xenium slides from the previous step were removed from −80 °C storage on dry ice prior to incubation at 37 °C for 1 min. Samples were then fixed in 4% paraformaldehyde (Formaldehyde 16% in aqueous solution, 100503-917, VWR) in PBS for 30 min. After rinsing in PBS, the samples were permeabilized in 1% SDS (sodium dodecyl sulfate solution) for 2 min, and then rinsed in PBS before being immersed in the pre-chilled 70% methanol and incubated for 60 min on ice. After rinsing the samples in PBS, the Xenium Cassettes were assembled on the slides. Samples were incubated with a probe hybridization mix containing both the Xenium Human Brain Gene Expression Panel (1000599, 10x Genomics) and a 100 custom gene panel at 50°C overnight to allow the probes to hybridize to targeted mRNAs. After probe hybridization, samples were rinsed with PBST, and incubated with Xenium Post Hybridization Wash Buffer at 37°C for 30 min. Samples were then rinsed with PBST and ligation mix was added. Ligation was performed at 37°C for 2 hrs to circularize the hybridized probes. After rinsing the samples with PBST, Amplification Master Mix was added to enzymatically amplify the circularized probes at 30 °C for 2 hrs. After washing with TE buffer, auto-fluorescence was quenched according to the manufacturer’s protocol and nuclei stained with DAPI prior to Xenium *in situ* analysis.

#### Data processing

The prepared samples were loaded into the Xenium analyzer and run according to manufacturer’s instructions “*Xenium Analyzer User Guide CG000584 Rev B*”. After the Xenium analyzer was initiated, the correct gene panel was chosen, and decoding consumables (Xenium Decoding Consumables, PN-1000487, 10x Genomics) and reagents (Xenium Decoding Reagents, PN-1000461, 10x Genomics) were loaded. The bottom of the slides were carefully cleaned with ethanol prior to loading. Once the samples were loaded and the run was initiated, the instrument scanned the whole sample area of the slides using the DAPI channel, and regions of interest were selected to maximize the capture area. Results were generated by the instrument using default settings. By default, the Xenium analyzer uses 15 µm nuclei expansion distance for segmentation of cells. To test the idea of nuclei only segmentation, we resegment the results with 0 µm nuclei expansion, by using the Xenium ranger and the following scripts:

~~~
xeniumranger resegment --id=demo --xenium-bundle=/path/to/xenium/files -- expansion-distance=0 --resegment-nuclei=True
~~~

#### Major cell type identification

After nuclei were segmented, cell x gene matrices were generated from the overlap of each segmented nuclear boundary with detected transcripts in the Xenium experiment. Nuclei were subsequently filtered by the number of detected transcripts, and only those containing at least 40 nuclear transcripts were retained for downstream analysis. Gene expression data from each sample was then log-normalized and normalized data were used for PCA, kNN graph calculation and Leiden clustering. Clusters were then assigned to one of 8 major cell types based on marker gene expression.

#### Label prediction from snRNA-seq data using scANVI

As an alternative to marker-based annotation, we used scANVI^119^ to perform reference-based label transfer from the RADC dataset. Briefly, we followed the following steps. First, snRNA-seq gene expression data was subset to the genes shared with the Xenium gene panel. Next, we used the scvi-tools package^120,121^ to train machine learning models for dimensionality reduction based on the reference dataset and its assigned labels (e.g., class and subclass). Models were run with 5 layers and 30 latent variables, and the scANVI model was trained for 20 epochs with a minimal sample of 100 cells per cluster per epoch. Lastly, a transfer model was trained for 100 epochs and applied to query data to assign labels based on those the model was trained on from the reference. To assess the performance of each transfer model, we asked the model to predict labels in the reference data (using the subset gene pool) and evaluated the rate of correct prediction and biases in label misassignment for each predicted label.

#### Subclass label transfer for EN (IN) cells

To assign subclass labels for EN and IN nuclei, nuclei from all samples were filtered to EN (IN) based on two alternative methods for major cell type prediction - that described above, and label transfer using scANVI (with the RADC dataset as a reference). After subsetting to nuclei labeled as EN (IN) by both methods, these were then used as a query for a second scANVI label transfer - this one trained on subclass labels. Accuracy for both major cell type and subclass models assessed by predicted labels in the RADC dataset based only on the Xenium gene panel was estimated at > 98%.

### Processing of genotypes

DNA extraction and genotyping was performed as described previously^122^. In brief, genomic DNA was extracted from frozen brain tissue using the QIAamp DNA Mini Kit (Qiagen), according to the manufacturer’s instructions. Samples were genotyped using the Infinium Psych Chip Array (Illumina) at the Mount Sinai Sequencing Core. Pre-imputation processing consisted of running the quality control script HRC-1000G-check-bim.pl from the McCarthy Lab Group (https://www.well.ox.ac.uk/~wrayner/tools/), using the Trans-Omics for Precision Medicine (TOPMed)^123^. Genotypes were then phased and imputed on the TOPMed Imputation Server (https://imputation.biodatacatalyst.nhlbi.nih.gov). Samples with a mismatch between one’s self-reported and genetically inferred sex, suspected sex chromosome aneuploidies, high relatedness as defined by the KING kinship coefficient^124^ (KING > 0.177), and outlier heterozygosity (+/− 3SD from mean) were removed. Additionally, samples with a sample-level missingness > 0.05 were removed and calculated within a subset of high-quality variants (variant-level missingness ≤ 0.02).

For ancestry assignment, genotypes were first merged with GRCh38 v2a 1000 Genomes Project data (https://wellcomeopenresearch.org/articles/4-50)^125^ using BCFtools version 1.9^126^. PLINK 2.0^127^ was then used to calculate the merged genotypes’ principal components (PCs), following filtering (minor allele frequency (MAF) ≥ 0.01, Hardy-Weinberg equilibrium (HWE) P ≥ 1 × 10^−10^, variant-level missingness ≤ 0.01, regions with high linkage disequilibrium (LD) removed) and LD pruning (window size = 1000 kb, step size = 10, r2 = 0.2) steps. For the samples of EUR ancestry assigned using the QDA method, autosomal bi-allelic variants with an imputation R^2^ > 0.8, HWE P ≥ 1 × 10^−6^, and variant-level missingness ≤ 0.02 were retained. Genotypes were then annotated with ancestry-specific MAF values from the National Center for Biotechnology Information’s Allele Frequency Aggregator (ALFA) (https://ftp.ncbi.nih.gov/snp/population_frequency/latest_release/). Only variants with an ancestry-specific ALFA MAF ≥ 0.01 were retained.

### Polygenic risk score calculation

Polygenic risk scores (PRS) were computed for the PsychAD cohort using summary statistics from AD GWAS^11^. The PRS-CS-auto method^128^ was employed, which incorporates continuous shrinkage priors to adjust the effect sizes from these summary statistics. An LD reference panel from the developers of PRS-CS, based on data from the 1000 Genomes Project^125^, was used (https://github.com/getian107/PRScs). The default settings for PRS-CS were applied, including parameters a = 1 and b = 0.5 for the γ-γ prior, 1000 Markov Chain Monte Carlo (MCMC) iterations, 500 burn-in iterations, and a thinning factor of 5. The global shrinkage parameter phi was determined using a fully Bayesian method. PLINK 2.0^127^ was utilized to calculate the individual-level PRS.

### Genetic heritability analysis of polygenic risk

We established a standardized pipeline for Multi-marker Analysis of GenoMic Annotation (MAGMA) followed by single-cell Disease-Relevance Scoring (scDRS). MAGMA incorporates the association P-values of genetic variants from the latest genome-wide association study (GWAS). We used the following GWAS summary stats in scDRS/MAGMA pipeline: AD^11^, MS^12^, PD^13^, Epilepsy^14^, Migraines^15^, Stroke^16^, ALS^17^, SCZ^18^, BD^19^, MDD^20^, ASD^21^, ADHD^22^, Insomnia^23^, Education^24^, IQ^25^, Alcoholism^26^, OCD^27^, Tourettes^28^, Obesity^29^, T2D^30^, Cholesterol^31^, RArthritis^32^, IBD^33^, UC^34^. We applied MAGMA using a standard window of 35 kbp upstream and 10 kbp downstream around the gene body. We executed scDRS using the top 1000 gene weights, sorted by Z score. The MAGMA and scDRS pipeline was run using the following parameters. MAGMA was run using `-snp-loc g1000_eur.bim` (SNP location file corresponding to the Phase 3 1000 Genome Project) and `--gene-loc NCBI38.gene.loc` (gene location file from NCBI build 38). Both files were obtained from https://ctg.cncr.nl/software/magma. For `scDRS`, default setting was applied.

### Variance partition analysis of gene expression

After aggregating pseudobulk by library, assays (cell types) were stacked using the *StackedAssay* function of Dreamlet. The resulting pseudobulk allowed us to perform analysis across cell types. Variance partition analysis was performed on the resulting stacked pseudobulk. We used the following regression formula:

~~~
Gene expression ∼ (1|stackedAssay) + (1|Channel) + (1|SubID) + (1|Source) + (1|Ethnicity) + dx_bit + scale(Age) + Sex + scale(PMI) + log(n_genes) + percent_mito + mito_genes + ribo_genes + mito_ribo
~~~

where *dx_bit* indicates binary disease status excluding metabolic and eating disorders. Technical covariates *log(n_genes)*, *percent_mito*, *mito_genes*, *ribo_genes*, and *mito_ribo* were removed from the plotting and subsequent analysis because they explained less than 1e-4 percent of overall gene expression variation.

### MAPT locus haplotyping

From our harmonized genotype calls, we selected common variants in 17q21.31 locus (chr17:45307631-46836264), performed PCA analysis of genotypes using 10 PCs, and used K-means clustering with k=3 to call three genotype clusters, H1H1, H1H2, and H2H2. We additionally confirmed the H1 haplotype using two published SNPs, rs17763050 and rs8070723, known to associate^129^. Haplotypes were estimated using Beagle v5.4^130^ on the selected genotypes of the 17q21.31 region. The estimation of the initial haplotype frequency model converged after one burn-in iteration, and the estimate of the genotype phase converged after 23 phasing iterations. For testing association with PD diagnosis, we used logistic regression with age, sex, 10 genotype PCs, and H1H1 status:

~~~
PD ∼ Age + Sex + Source + PC1 + PC2 + PC3 + PC4 + PC5 + PC6 + PC7 + PC8 + PC9 + PC10 + H1H1
~~~

In addition, we tested the contribution of the H1 haplotype to AD among non-ApoE4 carriers^49^ but did not find a significant association (P ≤ 0.302). For testing association with AD diagnosis, we first subsetted for individuals who are not carriers of the ApoE4 allele and tested for AD association using logistic regression with the formula:

~~~
AD ∼ Age + Sex + Source + PC1 + PC2 + PC3 + PC4 + PC5 + PC6 + PC7 + PC8 + PC9 + PC10 + H1H1
~~~

### Compositional variation analysis using Crumblr

We applied the Crumblr method (https://diseaseneurogenomics.github.io/crumblr) for testing the variation of cell type composition^131^. In summary, Crumblr scales the cell count ratio (i.e., fractions) data using centered log-ratio (CLR) transformation and applies linear models. Since CLR-transformed data is still highly heteroskedastic, the precision of measurements varies widely. Crumblr uses a fast asymptotic normal approximation of CLR-transformed counts from a Dirichlet-multinomial distribution to model the sampling variance of the transformed counts. Crumblr enables incorporating the sampling variance as precision weights to linear (mixed) models in order to increase power and control the false positive rate. Crumblr also uses a variance stabilizing transform based on the precision weights to improve the performance of PCA and clustering. Hypothesis testing was computed using the following formula:

~~~
Cell composition ∼ scale(Age) + Sex + (phenotype of interest)
~~~

By including these variables, we account for potential confounders and improve the accuracy and reliability of our hypothesis testing (**Supplementary Fig. 4b**).

### Differential gene expression analysis using Dreamlet

Due to the increased variable complexity in a large-scale disease atlas, scaling single-cell based approaches to millions of cells across a wide range of phenotypes presents computational challenges^132^ and can be suboptimal^133–136^. To account for the scale of these data, complex study designs with repeated measures, and low read count per cell, we applied Dreamlet for differential expression analysis, which applies a pseudobulk approach. Building from the previously developed statistical tool Dream^137^, it applies linear mixed models to the differential expression problem in single-cell omics data. It starts by aggregating cells by the donor using a pseudobulk approach^133,134^ and fits a regression model and cell. For each feature and cell cluster, the following mixed model was applied:

~~~
Gene expression ∼ scale(Age) + Sex + scale(PMI) + log(n_genes) + percent_mito + mito_genes + mito_ribo + ribo_genes + (phenotype of interest)
~~~

where categorical and numerical variables were modeled as random and fixed effects, respectively. If the phenotype of interest was a categorical variable, we set the intercept as 0 and used pre-defined contrasts between two factors. We ran a gene set analysis using the full spectrum of gene-level t-statistics^138^.

### Meta-Analysis (between brain sources)

We conducted a meta-analysis to integrate results from different brain banks for the same disorder. Data tables from multiple brain banks were combined into a single list for each disorder and annotated with their respective sources. The meta_analysis function was used to perform the meta-analysis, which involved combining data tables into a single data frame, grouping the data by assay, and calculating standard errors using the formula `abs(logFC / t)`. The meta-analysis was performed using the `rma` function from the `metafor` package with a fixed effects model. P-values were adjusted using the False Discovery Rate (FDR) method, and the negative log10 of the FDR values were calculated. This method was applied to datasets of AD, DLBD, Vas, PD, Tau, FTD, SCZ, and BD.

### Meta-Analysis (across the same disease category)

To further synthesize findings across multiple disorders, a `meta-of-meta` analysis was conducted, grouping the disorders into neurodegenerative and neuropsychiatric categories. Results from the initial meta-analyses for each disorder were combined into lists based on their categories. The `meta-of-meta` function was used to perform this higher-level analysis. This function combined the meta-analysis results into a single data frame, grouped the data by assay, and calculated standard errors using the formula `abs(estimate / statistic)`. This approach was applied to create `meta-of-meta` analyses for all disorders, neurodegenerative disorders, and neuropsychiatric disorders.

### Evaluation of shared disease signatures using Mashr

Suppose the total disease signature can be written as the sum of shared and distinct components of DEGs for that disease:

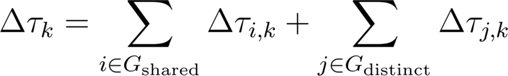

where *Δ_τk_* denotes the total disease signature for a disease *k*, *G_shared_* denotes the set of DEGs that are shared among the diseases, *G_distinct_* denotes the set of DEGs that are distinct for diseases. This notation highlights that the total differential gene expression for each disease is composed of contributions from both shared genes (common to all diseases) and distinct genes (specific to each disease). To define shared disease signatures, we performed composite tests on Dreamlet results using Mashr to evaluate the specificity of an effect. We evaluated the posterior probability of a non-zero effect present in all 8 cross-disorder contrasts. We categorized a gene as part of the shared component if it had Mashr posterior probability greater than 0.01.

### Construction of Correlation Matrix

To calculate the correlation matrix, we used a systematic approach to quantify the relationships between genetic estimates across different neuropsychiatric and neurodegenerative disorders. Spearman correlation coefficients were calculated to assess the strength and direction of association between genetic estimates across different disorders. The calculation was performed for the common to each pair of disorders, grouped by assay, after the exclusion of the shared disease signatures.

The wide-format correlation matrix was converted to a matrix suitable for heatmap visualization. Missing values were replaced with zeros. Annotations indicating the number of significant genes were added to the rows and columns using the `rowAnnotation` and `HeatmapAnnotation` functions from the `ComplexHeatmap` package. The heatmap was generated with hierarchical clustering of both rows and columns, and colored based on the Spearman correlation values using a gradient from blue (negative correlation) to red (positive correlation).

### Co-heritability analysis

We employed cross-trait LD score regression using the LDSC tool^139^ to estimate the genetic correlation between a pair of traits. We used summary stats for the following GWAS traits (AD^11^, DLBD^140^, PD^13^, FTD^141^, SCZ^18^, BD^19^) and calculated heritability for each of the traits and the genetic covariance and correlation between each of the pair of traits (in total 15 pairs of traits). The size of the cohort was provided to the function munge_sumstats.py for heritability estimates. Precomputed LD scores for 1000 Genomes EUR data were downloaded from https://data.broadinstitute.org/alkesgroup/LDSCORE/eur_w_ld_chr.tar.bz2. The SNP list for `munge_sumstats.py` was downloaded from https://data.broadinstitute.org/alkesgroup/LDSCORE/w_hm3.snplist.bz2. Standard error was obtained from the LDSC output. Script `munge_sumstats.py` was modified to include the parameter: `--chunksize 5e5`.

For each of the 15 possible combinations of traits, we calculated the level of correlation of gene expression using the Spearman rank correlation test. Genes were selected by applying the following criteria: logFC ≥ 0.5, FDR ≥ 0.05. Co-expression coefficient was calculated for the overall dataset and for each of the cell types. Next, to correlate co-expression and co-heritability, we calculated Spearman rank correlation coefficient between LDSC genetic correlation score and co-expression coefficient using 15 possible combinations of traits as data points for the Spearman rank correlation. Spearman was calculated for the overall dataset providing the genetic estimates of the expression similarities in the PsychAD cohort, and also per cell type to obtain the ranking of the cell types that contribute the most to the genetic to transcriptomic similarity in PsychAD.

### Mediation analysis

Causal Mediation Analysis was performed using two R packages; mediation and psych. Results were cross-validated between the two methods (identical within a threshold) to ensure the estimated coefficients, and the mediation effects are statistically robust. From 696 individuals in AD contrast, we subsetted 645 individuals with European ancestry who have PRS calculations from the latest AD GWAS^11^. For each regression, we used the following covariates:

~~~
Age + Sex + PMI + PC1 + PC2 + PC3 + PC4 + PC5 + PC6 + PC7 + PC8 + PC9 + PC10
~~~

where PC1-10 indicate genotype PCs. For subclass proportion, we used CLR-transformed cell count fractions from crumblr analysis. For bootstrapping, we used 10,000 simulations with 50th percentile of the treatment variable used as the control condition and 90th percentile of the treatment variable used as the treatment condition.

### Trajectory analysis using VAE model

#### Rationale

Traditionally, changes in gene expression as a function of disease state are measured using linear-based models. This approach has proven highly valuable and has enhanced our understanding of the biological mechanisms underlying many diseases. However, it is increasingly recognized that changes in gene expression can be highly nonlinear^60^; the interaction among numerous signaling pathways, many involving multiple feedback loops, can lead to complex dynamics that linear models may fail to capture. One approach to capturing potentially nonlinear changes in gene expression is pseudotime analysis (i.e., trajectory inference)^142,143^. In this approach, cells are assigned a relative pseudotime based on a metric that measures the distance between gene expression vectors, coupled with specific assumptions about how trajectories can evolve. However, this approach requires prior assumptions that might obscure informative aspects of disease progression. For example, most methods require a dimensionality reduction operation (e.g., PCA or UMAP) before calculating each cell’s nearest neighbors. The variance of lowly expressed genes, regardless of their importance to the disease, can be overshadowed by the variance of genes with greater expression but less relevance, or by the variance inherent in a sample from a highly diverse demographic. Furthermore, in the case of AD, it is known that the spread of neurofibrillary tangles and Aβ plaques is strongly, but not perfectly, correlated with dementia. Of special interest is understanding cases where individuals are resilient to dementia despite a high neurofibrillary tangle or Aβ plaque burden. Thus, we aimed to disentangle changes in gene expression associated with disease burden from those associated with the onset of dementia. Standard pseudotime analysis does not allow us to separate these two covariates effectively. To capture the potentially nonlinear gene expression dynamics during the course of AD progression and to disentangle the effects of disease burden and dementia, we used an alternative approach. As detailed below, we employed a VAE to predict the Braak stage and dementia status from the raw transcript counts of single cells. The model’s predictions of the Braak stage and dementia status were then used as two independent pseudotime axes. Importantly, we trained the model by equally sampling all combinations of Braak stage and dementia status, thereby discouraging the model from learning spurious correlations between the two target variables.

#### Model architecture

We used a VAE, based on the scVI model^144^, to predict the Braak stage and dementia status from single-cell gene counts. Both the gene encoder and decoder contained two 512-dimensional hidden layers. All hidden layers applied the ReLU activation function before a LayerNorm operation. Model predictions for Braak and dementia were generated using linear functions from the 32-dimensional latent layer.

#### Model training

Single-cell gene counts were log1p transformed in the input layer, and top 25,000 genes for each cell class, based on the percentage of cells the gene was expressed, were used for training. Genes found in the X or Y chromosomes were excluded to discourage the model from learning sex specific differences. The network was trained to minimize 1) the gene reconstruction loss, 2) the Kullback-Leiber divergence, and 3) the disease target prediction (Braak and dementia) loss:

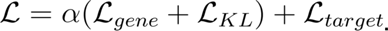

The scalar α was calibrated to properly weigh the contribution of the standard VAE loss terms (i.e., the gene reconstructions and KL losses) and the target and covariate prediction loss (see below). The gene reconstruction loss, *L_gene_*, was the zero-inflated negative binomial^144^. For the binary target dementia, the softmax function was applied to the output, and the loss was the cross-entropy. For Braak, target values were first normalized to zero mean and unit standard deviation, and the mean-squared error loss was used. All loss terms were trained simultaneously. To prevent the model from overfitting the data, we applied a dropout with 0.25 probability in the input layer (after the log1p transform) and with probability of 0.5 to all hidden layers (after the ReLU and LayerNorm operations). For each cell class, we divided the cells into 10 splits. In each split, 90% of the cells were used for training, and the remaining 10% were used for inference. Within each split, cells from a single donor exclusively belonged to either the training set or the inference set, but never both. Thus, model predictions were always based on cross-validated data from different donors. We trained one model for each of the ten splits to generate predictions for all donors for that cell class. Models were trained using all donors from the MSSM and RADC brain sources, and then ran inference on only the 696 donors that focused on the AD phenotype contrast (**Fig. 1d**). There existed different numbers of cells for each cell class, thus the amount of training differed between classes. For neurons, astrocytes, oligodendrocytes and the immune cell class, we trained for 5 epochs, and then calculated the Braak and dementia model predictions by averaging the inference outputs generated from the last two epochs. For mural and endothelial cells, which contained less data, we trained for 15 and 20 epochs, respectively, and calculated the Braak and dementia model predictions by averaging the outputs generated from the last five epochs. Model accuracy for the OPC cell class evolved more slowly over model training; thus, we also trained this cell class for 15 epochs, and calculated the Braak and dementia model predictions by averaging the outputs generated from the last five epochs. We used empirical testing to determine which values of *α* generated accurate Braak and dementia model predictions. Immune cells typically have reduced mean gene counts compared to the other cell classes, and we found that setting *α* = 0.005. For all other cell classes, in which mean gene counts were on average greater, we set *α* = 0.002. Network parameters were trained using the AdamW optimizer^145^ with parameters *β_2_* = 0.998, *ε* = 1*e* - 7) and weight decay of 0.05 applied to all layers before the latent layer. We used a batch size of 512, and a learning rate schedule with a warmup and decay period using the formula *lrate* = *d*^−0.05^ · *min* (*n*^−0.5^, *n* · *warmup*^−1.5^), where we set *d* = 5*e* - 4, *warmup* = 2000, while *n* is the current training step. Finally, we clipped the gradient norm to 0.25 to stabilize training.

#### Model accuracy

We averaged the cell-level Braak and dementia model predictions to obtain donor-averaged scores. For dementia, we calculated the balanced classification score by determining the percentage of donors without dementia with a prediction score ≤ 0.5, the percentage of donors with dementia with a prediction score > 0.5, and then averaging these two values. For Braak, we calculated the Pearson R value between the actual Braak stage and the Braak predictions. Error bars were generated using a bootstrap procedure, in which we randomly sampled donors (with replacement), calculated the Braak and dementia prediction accuracy, and repeated this process 20,000 times. For each cell class, we included all donors with at least 5 cells.

#### Disentangling Braak and dementia

Braak stage and dementia status are significantly correlated (Pearson R = 0.582, P < 1e-60). This strong correlation between target variables implies that input features (i.e., changes in gene expression) associated with the two target variables are also likely to be correlated, which makes it challenging for the model to learn which input feature is predictive of which target variable. The result is when the model is trained in a standard fashion, the correlation between the Braak and dementia model predictions are close to 1 (**Supplementary Fig. 7a**, Immune class shown), suggesting that the model has learned spurious correlations between the input features and target variables. Fully removing spurious correlation in machine learning models is still an unresolved question. However, balancing the training data, such that each of the 14 combinations of Braak and Dementia (7 Braak values X 2 Dementia values) are equally sampled, can effectively reduce spurious correlations learned during training^146^. In practice, we equally sampled from 15 groups, in which the extra group consisted of donors whose Braak stage or dementia status had not been determined. Training with group balancing reduced the correlation between the predicted Braak and dementia values (**Supplementary Fig. 7b)**, did not adversely affect the model accuracy at the donor-level (**Supplementary Fig. 7c**).

#### Calculating gene trajectories

We wished to measure how gene expression varied as a function of the predicted Braak stage. First, gene counts were normalized so that each cell’s total count was 10,000, followed by the log1p transformation. Second, for each cell class and each donor, we calculated the mean predicted Braak stage, and the mean normalized expression for each gene. Averaging within each donor reduced variability and ensured that donors with greater cell counts did not contribute disproportionately in downstream analysis. Third, we smoothed both the predicted donor-averaged Braak scores, and the donor-averaged gene expression with a Gaussian kernel (**Supplementary Fig. 8**). Specifically, for each donor *i*, we weighted all other donors *j* as

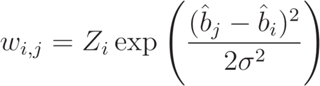

where 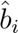 is the predicted Braak score of donor *i*, *σ*^2^ was set to half the variance of the predicted Braak distribution, and the normalization term *Z_i_* was set such that 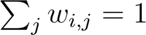. This allowed us to calculate smoothed Braak, 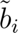 and gene expression, 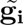, values

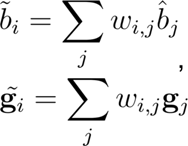

where g_i_ is the gene expression vector of log1p normalized counts for donor *i*. After ordering the smoothed Braak scores, Braak gene trajectories are now represented as the tuple

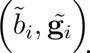

We only included donors with at least 5 cells for the cell class. We also removed the 10 donors with the least and greatest smoothed predicted Braak scores after smoothing to minimize the edge effects from smoothing.

#### Resilience against dementia

Since tau proteinopathy and dementia status are highly correlated, gene expression as a function of the two variables is also correlated, and thus partially redundant. Therefore, we aimed to measure how gene expression covaried with predicted dementia given the predicted Braak staging. To do so, we first calculated the expected predicted dementia and expected gene expression for donors with similar Braak staging. Specifically, we defined the expected dementia given Braak, 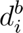, and the expected gene expression given Braak, 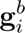,

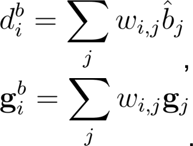

The only modification to the kernel was that we set *w_i,i_* = 0 so that each donor doesn’t contribute to its own expected value. We then calculated the residuals between the donor’s predicted dementia status and gene expression with its expected values:

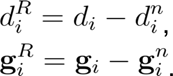

The dementia resilience score for each donor was then the product of these two terms. When calculating early and late resilience, donors were separated into early and late groups based on their predicted Braak staging before averaging within each group. Using this metric, we define genes as protective if gene expression increases as predicted dementia decreases, given the predicted Braak staging (i.e., the product of the terms defined above is negative). Conversely, we define genes as damaging if gene expression increases as predicted dementia increases, given the predicted Braak staging.

#### Trajectory nonlinearity

We were interested in measuring how gene expression potentially changes in a nonlinear manner as a function of Braak or dementia. As an initial step, we first wished to quantify the degree of nonlinearity in gene trajectories across the different cell classes. As outlined in **Supplementary Fig. 8**, this calculation was performed across a series of steps. Starting with the gene trajectories for each gene and each cell class as described above, in which smoothed gene expression varied as a function of predicted Braak, we fit each one with a linear fit, and then calculated its explained variance. We calculated the mean explained variance using all genes within each cell class with mean normalized expression above 0.01 and used this value as a baseline. Next, we fit each trajectory with a piecewise linear fit consisting of two consecutive linear segments, separated at some predicted Braak score 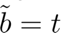. For each cell class, we determined the optimal *t* that maximized the explained variance of the piecewise fits for all genes within that cell class. The nonlinearity index (**Fig. 7b, bottom**) was then defined as the difference in explained variance between the piecewise linear fit and the linear fit. Error bars were determined by bootstrapping the model fits, in which we randomly sampled genes from donors (with replacement), calculating the nonlinearity index, and repeating the process 100 times. Significance between different cell classes was calculated by comparing the 100 x 100 bootstrapped nonlinear index values. We used the optimal time *t* was then used to define an “early” and “late” period. For the Immune cell class, which had the greatest nonlinearity index, the early period was defined as the first 140 donors with the lowest Braak model predictions (after the first 10 donors were removed to account for edge effects, see above). For the other cell classes, we adjusted 140 proportionally based on the number of donors for that cell class and defined this number as N; the early period for each cell class was then defined as the N donors with the lowest Braak model predictions. Since the rank ordering of donor-averaged Braak scores varies between cell classes, donors may be classified as early stage for some cell classes and late stage for others.

#### Trajectory gene enrichment

We wished to determine which genetic pathways were most significantly up or down-regulated during the progression of AD. To do so, we first extracted the slopes of the “early” and “late” linear fits for the Braak trajectories, and the mean early and late resilience scores (defined above). We used these slopes input to Zenith (https://bioconductor.org/packages/release/bioc/html/zenith.html) to calculate the changes across all GO BP pathways across the eight cell classes. For each GO BP pathway, we calculated the maximum - log10(FDR) score across the eight cell classes and across early and late Braak and Dementia stages. The score of each pathway was assigned this maximum value.

To obtain a condensed list of the most significantly genetic pathways, we first selected all pathways with FDR ≤ 0.05. Next, since we were only interested in pathways that could be informative of the mechanisms underlying AD progression, we excluded pathways containing words referring to overly broad behaviors or cognitive functions (“learning”, “memory”, “vocalization”, “social”, “auditory”, “startle response”, “behavior”, “locomotor”, “startle”, “prepulse inhibition”), terms referring to anatomical structures other than the cortex (“substantia nigra development”, “cardiac”, “coronary”, “aortic”, “ventricular”, “kidney”, “metanephric”, “retina”, “optic”, “bone”, “respiratory”, “pulmonary”, “olfactory”, “sperm”, “placenta”, “egg”, “embryonic”, “ovulation”, “estrous”, “placenta”, “sperm”, “mamary”, “germ layer”, “outflow tract septum”, “adrenal”, “epithelial”, “skeletal”, “otic”, “head”) or overly broad neural terms (“nervous system process”, “cerebral cortex”, “recognition”, “host”, “organ”, “developmental growth”). To condense the remaining pathways into a more manageable size, we used rrvgo (https://www.bioconductor.org/packages/release/bioc/html/rrvgo.html). We selected the Wang semantic similarity metric^147^, and set the threshold at 0.9 to obtain 56 GO BP pathways (**Supplementary Fig. 9**). For easier visualization, we selected 32 representative pathways from this set for **Fig 7c**. We also performed similar steps (pathways with minimum FDR < 0.05 included, Wang metric with threshold at 0.9) to obtain the top GO Molecular Function and Cellular Component (**Supplementary Figs. 10, 11**) pathways. In all cases, we only included pathways with at least 10 genes to ensure that results were statistically robust, and no more than 250 genes to ensure that pathways were not overly broad.

#### Mean trajectories and MAGMA enrichment

For both the mean normalized expression (**Fig. 7d**) and Magma enrichment analysis (**Fig. 7f**), we used the top 250 coding genes based upon the early and late slopes of the Braak trajectories. Late decreasing genes tend to also appear to contain an early increase (**Fig. 7d**). However, we cannot say whether this is biologically meaningful or the result of selection bias, as a strong late decrease must be preceded by a high baseline. For both the mean expression and Magma enrichment calculations, results were qualitatively similar if we used the top 500 or 1000 genes instead (data not shown).

## Data availability

All data are available via the AD Knowledge Portal (https://adknowledgeportal.org). The AD Knowledge Portal is a platform for accessing data, analyses, and tools generated by the Accelerating Medicines Partnership (AMP-AD) Target Discovery Program and other National Institute on Aging (NIA)-supported programs to enable open-science practices and accelerate translational learning. The data, analyses and tools are shared early in the research cycle without a publication embargo on secondary use. Data is available for general research use according to the following requirements for data access and data attribution (https://adknowledgeportal.synapse.org/Data%20Access). The data are available under controlled use conditions set by human privacy regulations. To access the data, a data use agreement is needed. The registration is in place solely to ensure the anonymity of the study participants. In addition, we have a data descriptor manuscript^102^ detailing the data processing and data collection.

## Code availability

All the source codes used in this study are available via GitHub https://github.com/DiseaseNeuroGenomics/PsychADxD.

## Acknowledgments

We would like to express our deep gratitude to the patients and their families who generously donated the invaluable biological material essential for the success of this study. We are profoundly indebted to their participation and commitment to advancing scientific knowledge and improving human health. We acknowledge the National Institute on Aging for their generous support in funding this research with the following NIH grants: R01AG067025, R01AG082185, and R01AG065582. Human tissues were obtained from the NIH NeuroBioBank at the Mount Sinai Brain Bank (MSSM; supported by NIMH-75N95019C00049), the Rush Alzheimer’s Disease Center (RADC; funding: P30AG10161, P30AG72975, R01AG15819, R01AG17917, R01AG22018, U01AG46152, and U01AG61356), and NIMH-IRP Human Brain Collection Core (HBCC, project # ZIC MH002903). This work was also supported by the Novo Nordisk Foundation NNF14CC0001 and NNF20SA0035590. The results published here are in whole or in part based on data obtained from the AD Knowledge Portal.

## Author contributions

Conceptualization: PR, VH, JFF, DL

Methodology: PR, JFF, DL, GEH

Software: GEH, DL, NYM

Validation: DL, XW, JMF, JFF, PK

Formal analysis: DL, MK, NYM, GEH, SK, MP, TC, FT

Investigation: JFF, AH, CC, ZS, MA, SA, XW, JMF

Resources: VH, DAB, SM, LLB, PKA

Data Curation: JB, PNM, DM, DB, KT, DL

Writing: DL, PR, JFF, NYM with supports from all co-authors.

Visualization: DL, MK, NYM

Supervision: PR, DL, JFF, JB, VH, GV, KG, LJJ

Project administration: DL, PR

Funding acquisition: PR, VH

All authors read and approved the final draft of the paper.

## Competing interests declaration

The authors declare no competing interests.

## Materials & Correspondence

Correspondence to Donghoon Lee or Panos Roussos.

## Supplementary Information

### PsychAD Consortium Authors

Aram Hong (1, 4, 6, 7); Athan Z. Li (10, 12); Biao Zeng (1, 4, 6, 7); Chenfeng He (9, 12); Chirag Gupta (9, 12); Christian Porras (1, 4, 6, 7); Clara Casey (1, 4, 6, 7); Colleen A. McClung (18); Collin Spencer (1, 4, 6, 7); Daifeng Wang (9, 10, 12); David A. Bennett (19); David Burstein (1, 2, 4, 6, 7, 8); Deepika Mathur (1, 4, 6, 7); Donghoon Lee (1, 4, 6, 7); Fotios Tsetsos (1, 2, 4, 6, 7); Gabriel E. Hoffman (1, 2, 4, 6, 7, 8); Genadi Ryan (13, 17); Georgios Voloudakis (1, 2, 3, 4, 6, 7, 8); Hui Yang (1, 4, 6, 7); Jaroslav Bendl (1, 4, 6, 7); Jerome J. Choi (11, 12); John F. Fullard (1, 4, 6, 7); Kalpana H. Arachchilage (9, 12); Karen Therrien (1, 4, 6, 7); Kiran Girdhar (1, 4, 6, 7); Lars J. Jensen (21); Lisa L. Barnes (19); Logan C. Dumitrescu (22, 23); Lyra Sheu (1, 4, 6, 7); Madeline R. Scott (18); Marcela Alvia (1, 4, 6, 7); Marios Anyfantakis (1, 4, 6, 7); Maxim Signaevsky (6, 7); Mikaela Koutrouli (1, 4, 6, 7, 21); Milos Pjanic (1, 4, 6, 7); Monika Ahirwar (13, 17); Nicolas Y. Masse (1, 4, 6, 7); Noah Cohen Kalafut (10, 12); Panos Roussos (1, 2, 4, 6, 7, 8); Pavan K. Auluck (20); Pavel Katsel (6); Pengfei Dong (1, 4, 6, 7); Pramod B. Chandrashekar (9, 12); Prashant N.M. (1, 4, 6, 7); Rachel Bercovitch (1, 4, 6, 7); Roman Kosoy (1, 4, 6, 7); Sanan Venkatesh (1, 2, 4, 6, 7); Saniya Khullar (9, 12); Sayali A. Alatkar (10, 12); Seon Kinrot (1, 4, 6, 7); Stathis Argyriou (1, 4, 6, 7); Stefano Marenco (20); Steven Finkbeiner (13, 14, 15, 16, 17); Steven P. Kleopoulos (1, 4, 6, 7); Tereza Clarence (1, 4, 6, 7); Timothy J. Hohman (22, 23); Ting Jin (9, 12); Vahram Haroutunian (5, 6, 7, 8); Vivek G. Ramaswamy (13, 17); Xiang Huang (12); Xinyi Wang (1, 4, 6, 7); Zhenyi Wu (1, 4, 6, 7); Zhiping Shao (1, 4, 6, 7)

### PsychAD Consortium Affiliations

1: Center for Disease Neurogenomics, Icahn School of Medicine at Mount Sinai, New York, NY, USA

2: Center for Precision Medicine and Translational Therapeutics, James J. Peters VA Medical Center, Bronx, NY, USA

3: Department of Artificial Intelligence and Human Health, Icahn School of Medicine at Mount Sinai, New York, NY, USA

4: Department of Genetics and Genomic Sciences, Icahn School of Medicine at Mount Sinai, New York, NY, USA

5: Department of Neuroscience, Icahn School of Medicine at Mount Sinai, New York, NY, USA

6: Department of Psychiatry, Icahn School of Medicine at Mount Sinai, New York, NY, USA

7: Friedman Brain Institute, Icahn School of Medicine at Mount Sinai, New York, NY, USA

8: Mental Illness Research, Education and Clinical Center VISN2, James J. Peters VA Medical Center, Bronx, NY, USA

9: Department of Biostatistics and Medical Informatics, University of Wisconsin-Madison, Madison, WI, USA

10: Department of Computer Sciences, University of Wisconsin-Madison, Madison, WI, USA

11: Department of Population Health Sciences, University of Wisconsin-Madison, Madison, WI, USA

12: Waisman Center, University of Wisconsin-Madison, Madison, WI, USA

13: Center for Systems and Therapeutics, Gladstone Institutes, San Francisco, CA, USA

14: Department of Neurology, University of California San Francisco, San Francisco, CA, USA

15: Department of Physiology, University of California San Francisco, San Francisco, CA, USA

16: Neuroscience and Biomedical Sciences Graduate Programs, University of California San Francisco, San Francisco, CA, USA

17: Taube/Koret Center for Neurodegenerative Disease Research, Gladstone Institutes, San Francisco, CA, USA

18: Department of Psychiatry, University of Pittsburgh School of Medicine, Pittsburgh, PA, USA

19: Rush Alzheimer’s Disease Center and Department of Neurological Sciences, Rush University Medical Center, Chicago, IL, USA

20: Human Brain Collection Core, National Institute of Mental Health-Intramural Research Program, Bethesda, MD, USA

21: Novo Nordisk Foundation Center for Protein Research, Faculty of Health and Medical Sciences, University of Copenhagen, Copenhagen, Denmark

22: Vanderbilt Genetics Institute, Vanderbilt University Medical Center, Nashville, TN, USA

23: Vanderbilt Memory & Alzheimer’s Center, Vanderbilt University Medical Center, Nashville, TN, USA

## Supplementary Figures

**Supplementary Fig. 1.**
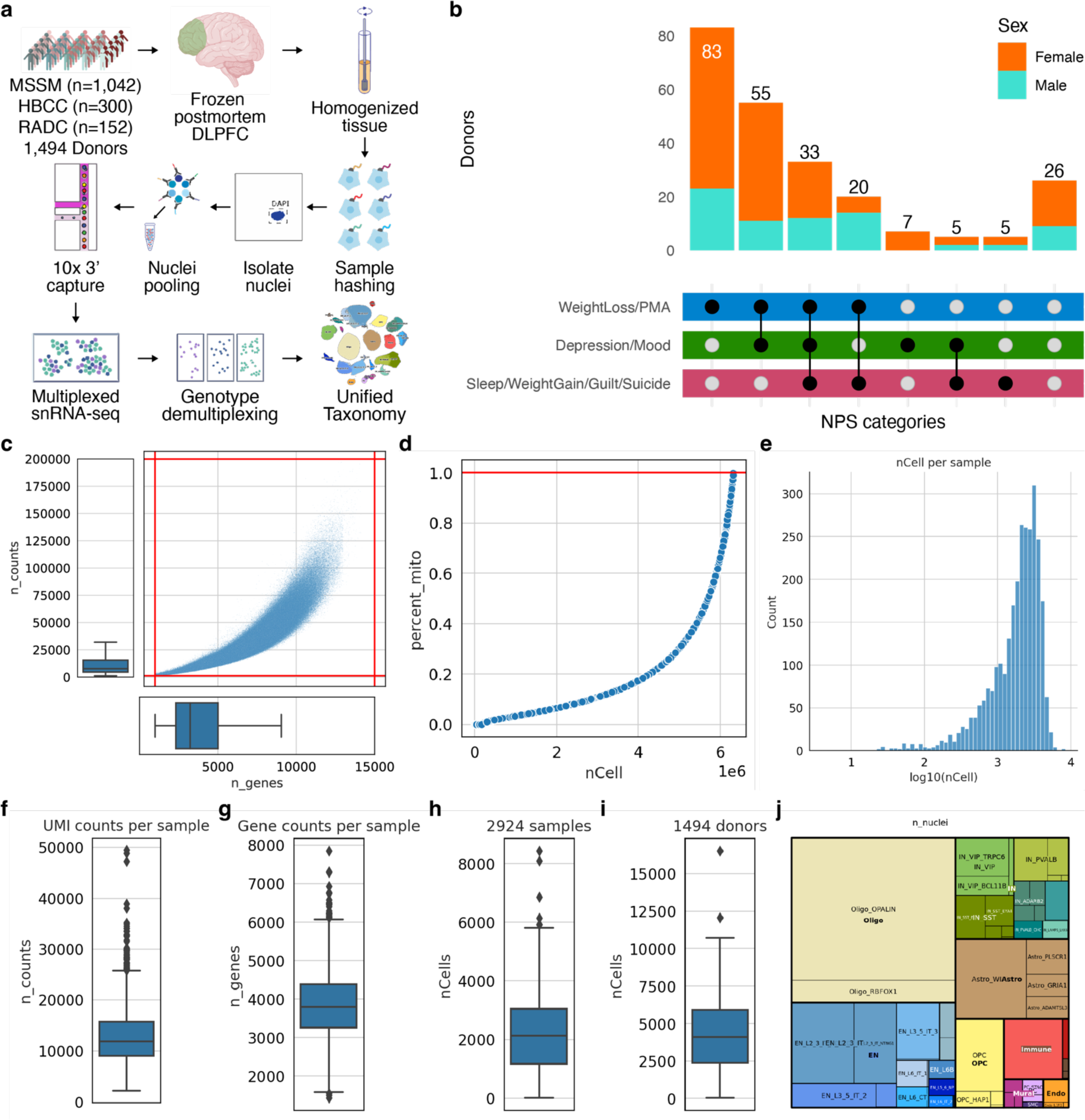
**(a)** Extended schematic overview of snRNA-seq experiments leading to unified taxonomy. **(b)** Donor breakdown by NPS categories and sex. **(c)** Distribution of gene counts by UMI counts after QC of 6M nuclei. **(d)** Cumulative nuclei count by percent mitochondrial genes. **(e)** Histogram of nuclei counts per sample. **(f)** Distribution of UMI counts per sample. **(g)** Distribution of gene counts per sample. **(h)** Distribution of nuclei count per sample. **(i)** Distribution of nuclei count per donor. **(j)** Treemap showing the color scheme of the unified taxonomy.

**Supplementary Fig. 2.**
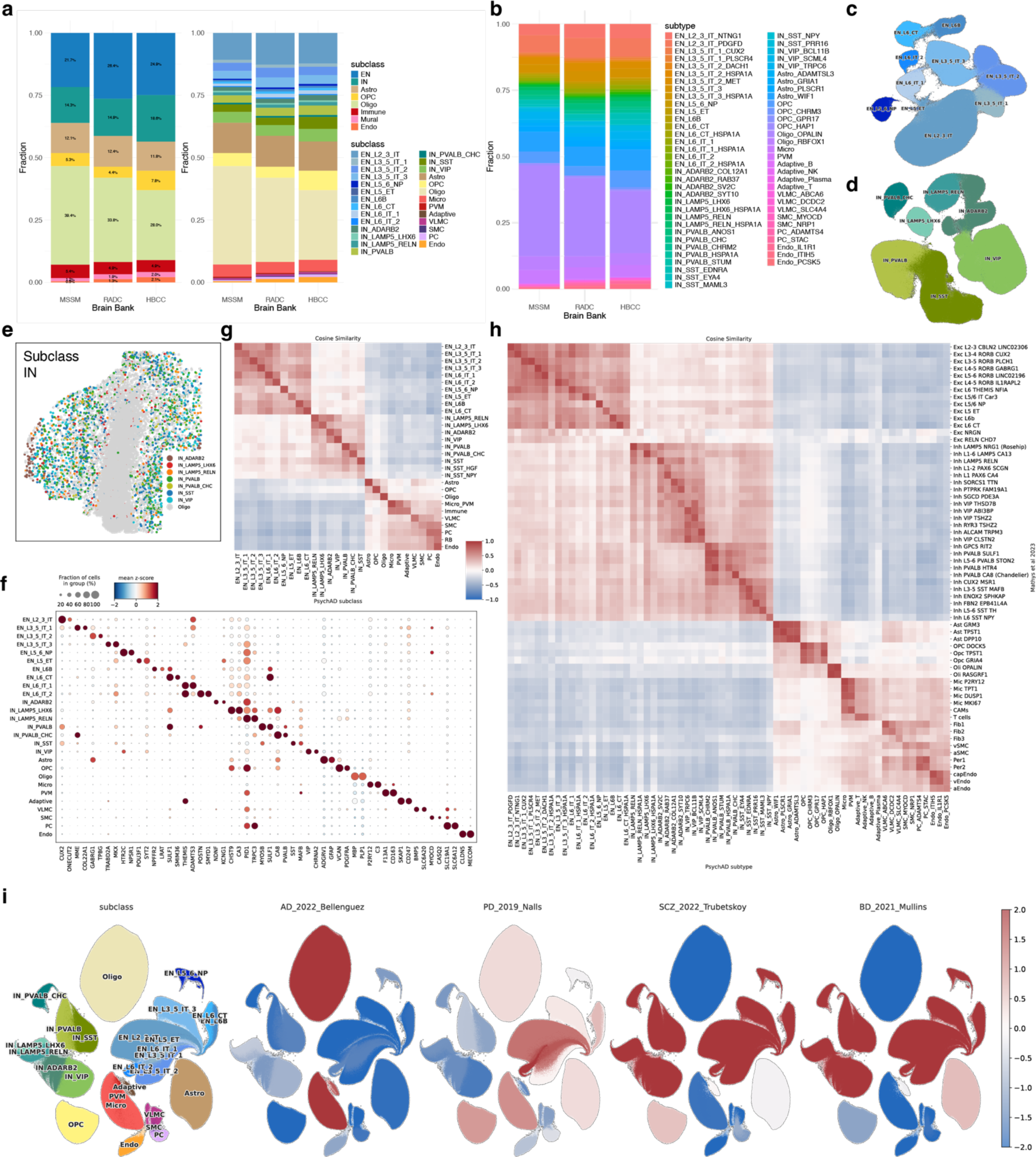
**(a)** Composition of cell types in each brain bank using class and subclass-level cellular taxonomy. **(b)** Composition of subtype in each brain bank. **(c)** Clustering of EN lineage. A UMAP focusing on the diversity of 10 subclasses of excitatory neurons. **(d)** Clustering of IN lineage. A UMAP focusing on the diversity of 7 subclasses of inhibitory neurons (IN). **(e)** Spatial distribution of IN subclass. **(f)** Expression and fraction of marker gene expression in 27 subclasses. Expression z-scaled and averaged across nuclei. **(g)** Comparison of subclass-level cellular taxonomy with Ma et al 2022. **(h)** Comparison of subtype-level cellular taxonomy with Mathys et al 2023. **(i)** Disease enrichment scores based on GWAS (scDRS) for AD, PD, SCZ, and BD.

**Supplementary Fig. 3.**
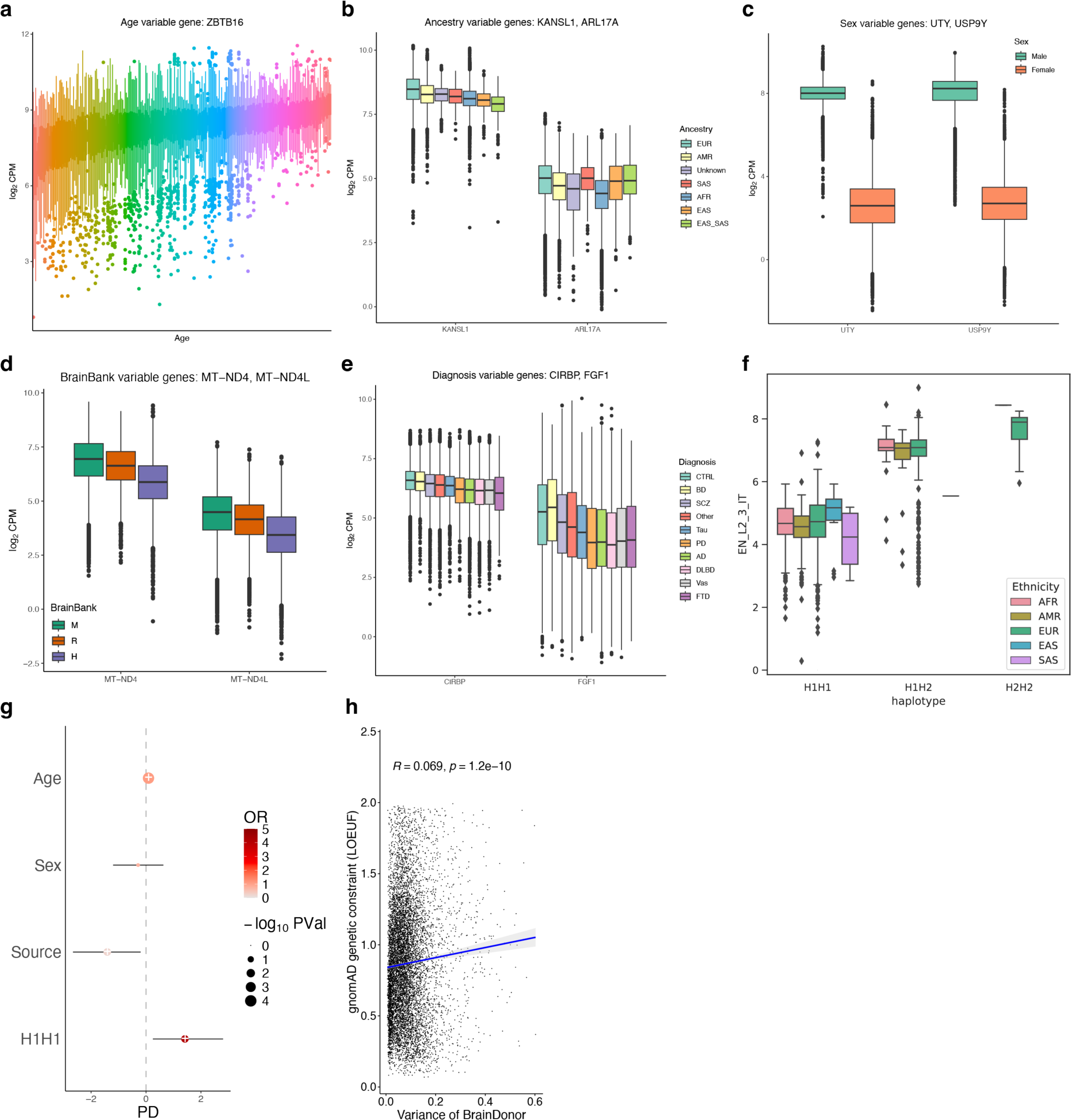
**(a)** Example of a gene with the highest variation across age. **(b)** Example of a gene with the highest variation across genetic ancestry. **(c)** Example of a gene with the highest variation across sex. **(d)** Example of a gene with the highest variation across brain banks. **(e)** Example of a gene with the highest variation across diagnosis. **(f)** Normalized expression of ARL17B gene within EN_L2_3_IT subclass stratified by genetic ancestry. **(g)** The logistic regression coefficient for PD risk using the haplotype of MAPT locus. **(h)** Spearman correlation test between variance explained by inter-individual variation and average LOEUF conservation score.

**Supplementary Fig. 4.**
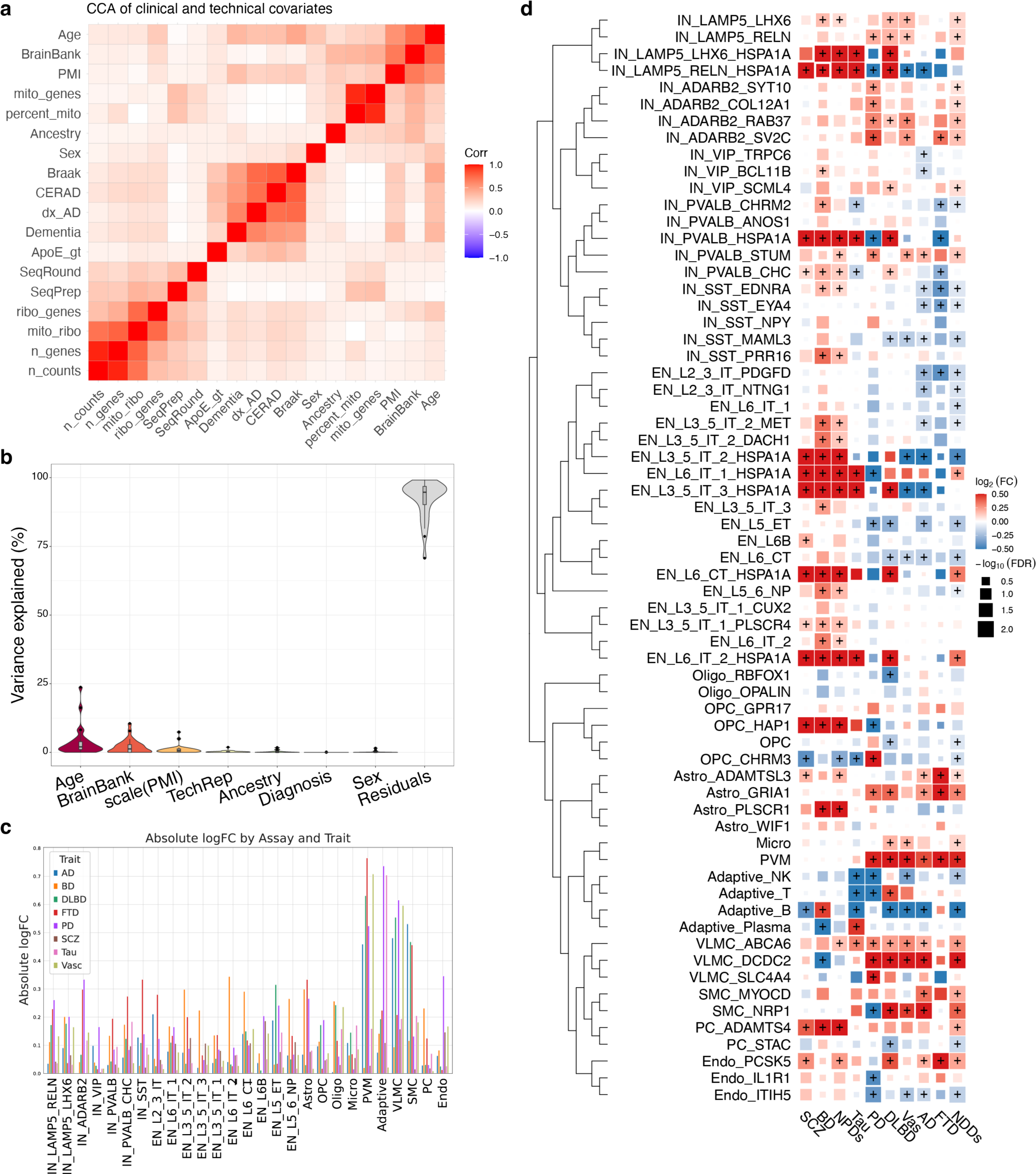
Cross-disorder variation of cell type composition comparing 8 different NDDs and NPDs against common neurotypical controls. **(a)** Correlation matrix showing the relationships between various clinical and demographic variables for the selection of the covariates. The color intensity indicates the strength of the correlation, with red representing positive correlations and blue negative correlations. **(b)** Variance partition of cell type composition, displaying the variance captured by different covariates in the formula. **(c)** Bar plot depicting the absolute logFC values of the 8 different NDDs and NPDs across the cell types. **(d)** Variation in cell type composition for each subtype in 8 different diseases. NDDs and NPDs indicate meta-analysis using broad disease categories. Color intensity indicates effect size and dot size reflects the statistical significance of correlations.

**Supplementary Fig. 5.**
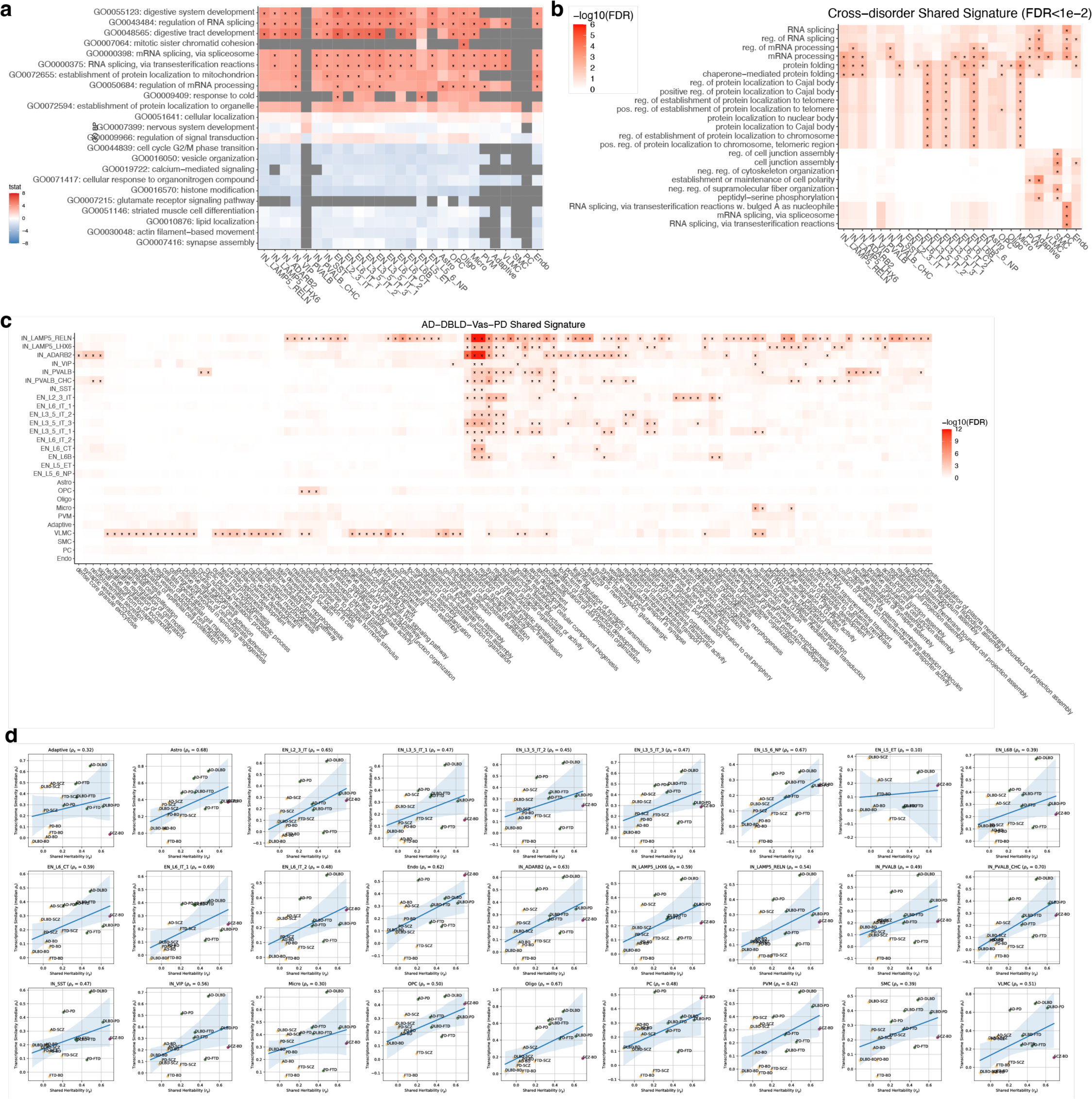
Cross-disorder gene expression signature. **(a)** GO BP pathways implicated by shared gene expression changes across 8 disorders using Zenith. T-statistic values indicate the direction and magnitude of the gene set enrichment. **(b)** Extended data for Fig. 5a with all 27 subclasses and without reduction in similar GO terms. **(c)** Pathways implicated by shared signatures from AD, DLBD, Vas, and PD. **(d)** Correlation between shared heritability and transcriptome similarity per cell subclass.

**Supplementary Fig. 6.**
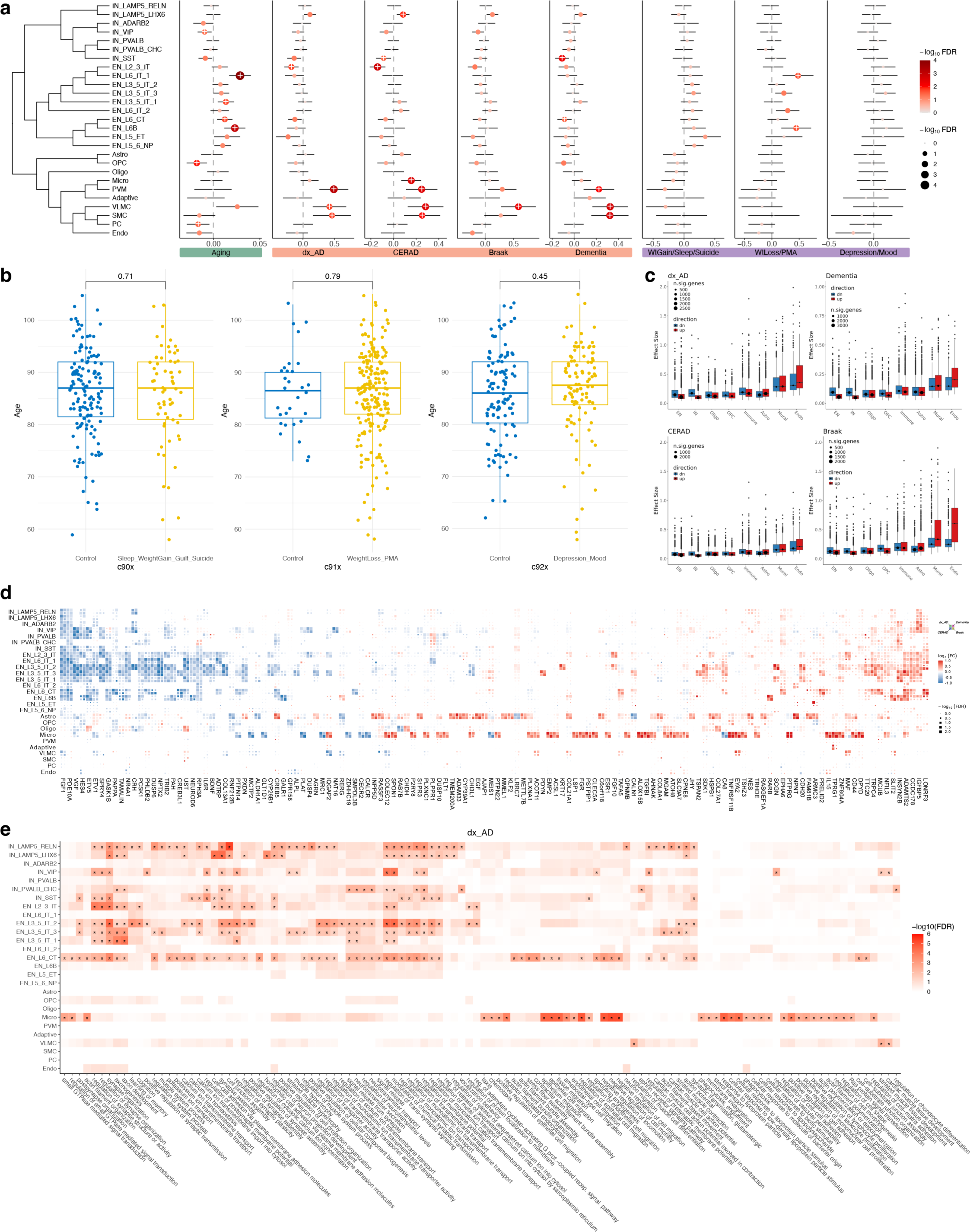
Transcriptomic changes across AD neuropathology. **(a)** Extended data for Fig. 6a. Compositional variation analysis using normal aging, different measures of AD pathology (binary AD diagnosis (dx_AD), CERAD score, Braak staging, and ordinal dementia scale), and 3 categories of NPS within AD. **(b)** Age distribution of AD cases with or without three NPS categories. **(c)** Mean effect sizes aggregated by direction of effect per cell class. **(d)** Extended data for Fig. 6d. DEGs in AD phenotypes. Meta-analysis between brain banks. Top genes with FDR < 0.01 and effect size ≥ 0.35. **(e)** Functional enrichment analysis of DEGs by subclass using Gene Ontology Biological Process. Hypergeometric test with FDR ≤ 0.01 shown.

**Supplementary Fig. 7.**
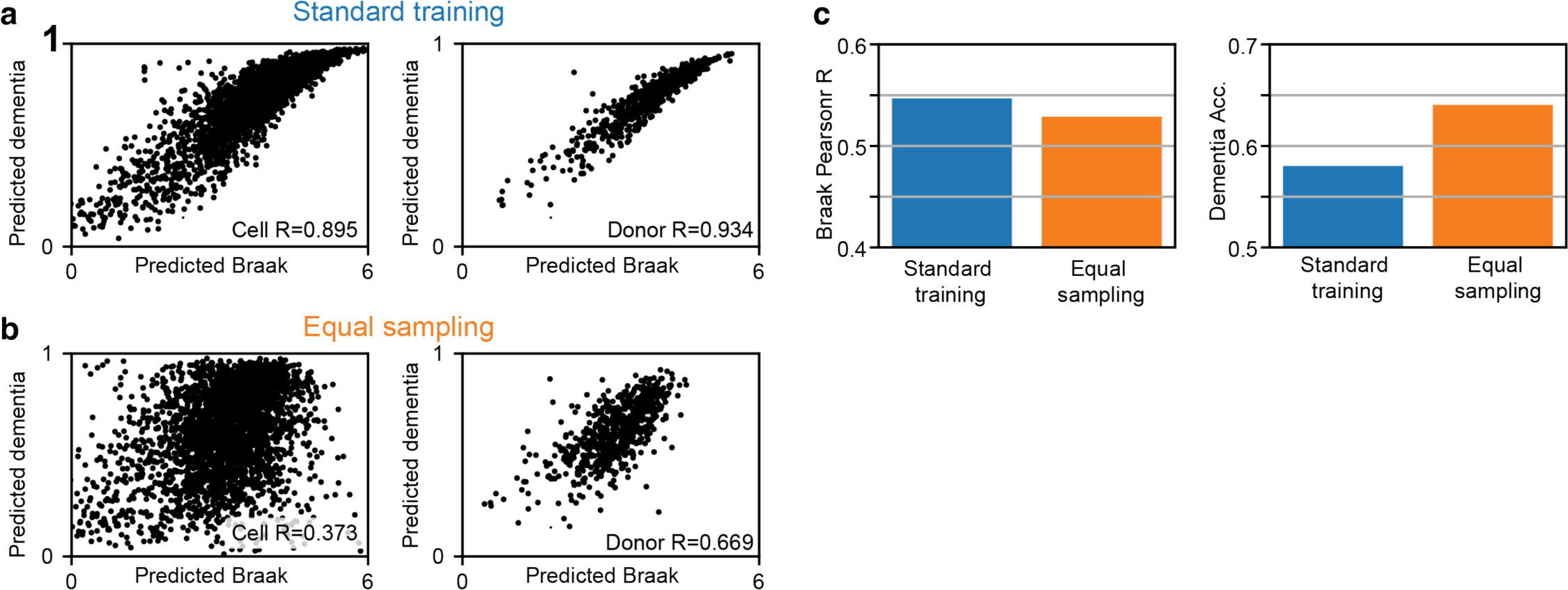
The effect of different training methods on model prediction correlations and accuracy for the Immune cell class. **(a)** Correlation between Braak (x-axis) and dementia (y-axis) model predictions at the cell-level (left panel) and donor-averaged level (right panel) when all cells are equally sampled. To reduce the figure size, only 1 out of 20 cells shown in the left panel. **(b)** Same as (a), except that all combinations of Braak stage and dementia status are equally sampled during training. This has the effect of reducing the correlation between the predicted Braak and dementia values. **(c)** The correlation between the actual and predicted Braak values (left panel) and dementia classification accuracy (right panel), both measured at the donor-averaged level, are shown for the two training methods.

**Supplementary Fig. 8.**
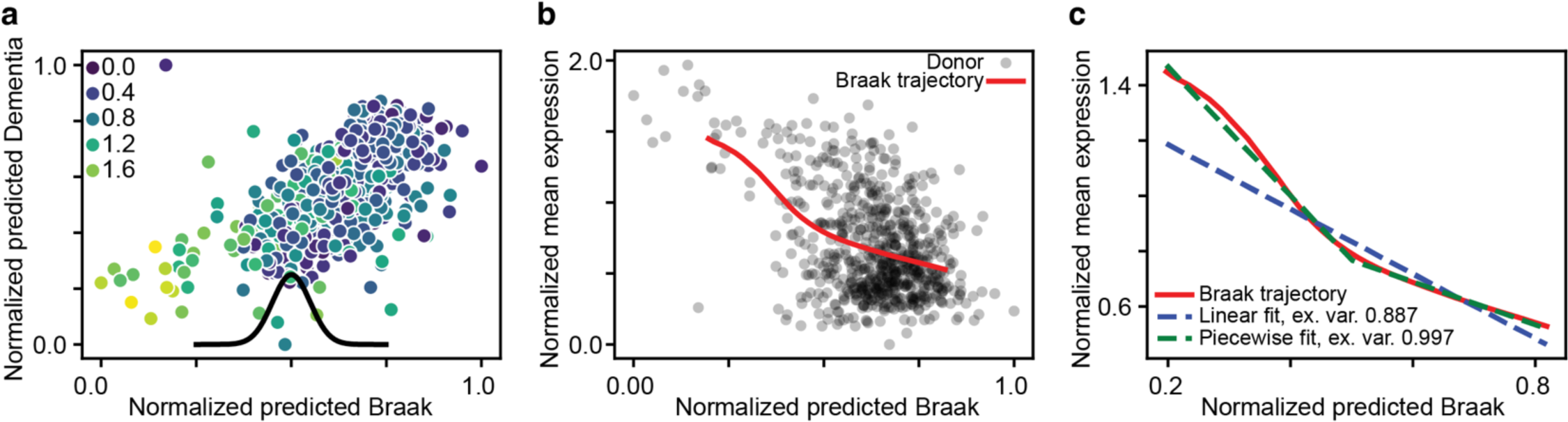
The steps involved in calculating the Braak trajectory for an example gene - NAV2 from the Immune cell class. **(a)** Given cell-based model predictions along with normalized gene counts, we can calculate the donor-averaged predicted Braak (x-axis), predicted dementia (y-axis), and normalized gene counts (hue). Predicted Braak and dementia values are normalized between 0 and 1. We then smooth the predicted Braak scores along with the gene counts with a Gaussian kernel (black curve, width is shown at scale). **(b)** After smoothing, we obtain the Braak trajectory for this gene, relating gene expression to the predicted Braak value. **(c)** We then compare how well we can fit this trajectory with a single linear model (blue dashed line) and a piecewise linear model (green dashed line). The piecewise linear model consists of two consecutive linear fits, in which the transition point has been optimized to maximize the explained variance. This transition point is then used to define an “early” and “late” disease stages (**Methods**).

**Supplementary Fig. 9.**
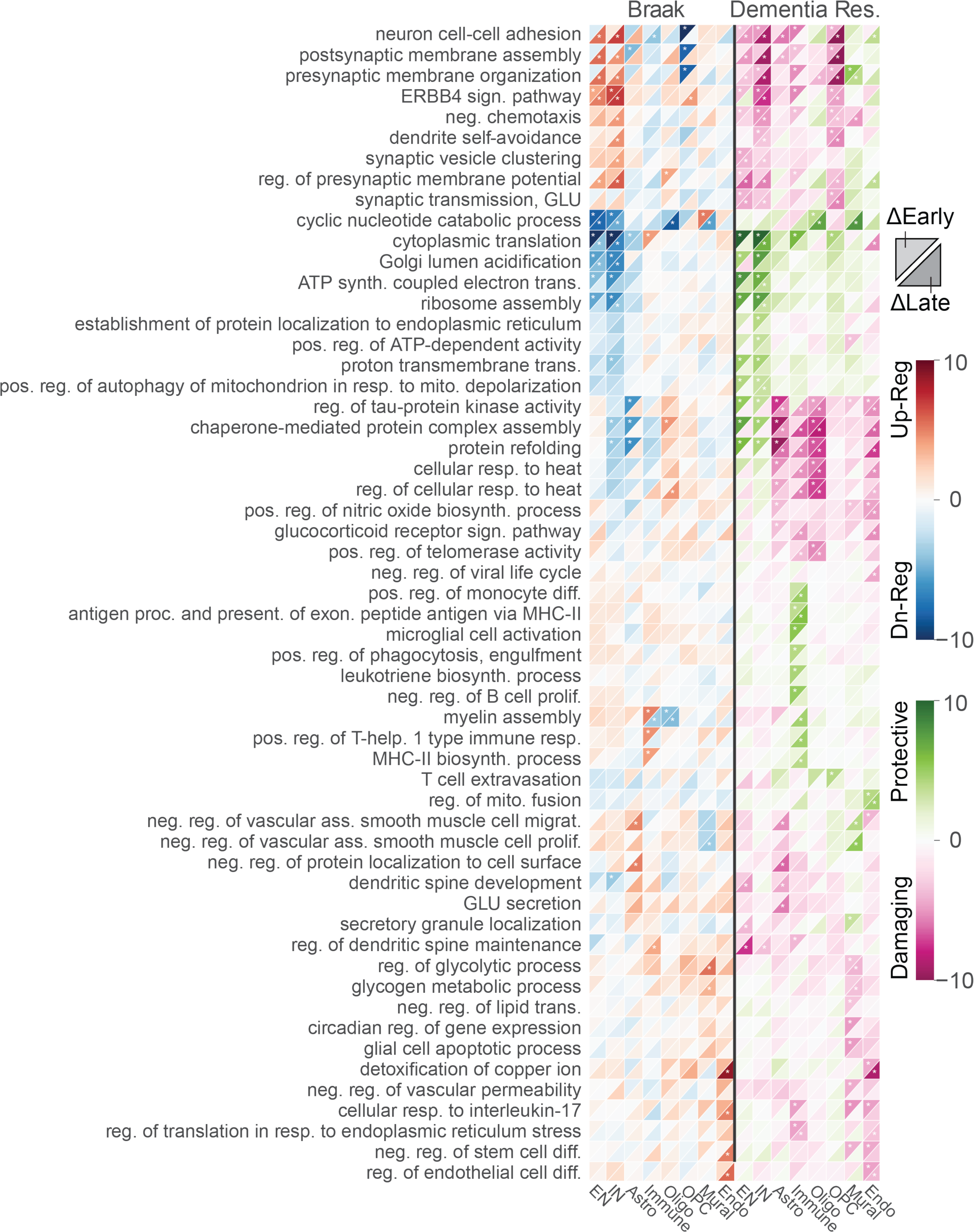
Extended data for Fig. 7c. Expanded list of all 56 GO BP pathways. Upper left triangles indicate the early phases, and lower right triangles indicate the late phases of the AD trajectory predicted by Braak and dementia resilience. Hue indicates the z-score, and stars indicate FDR < 0.05.

**Supplementary Fig. 10.**
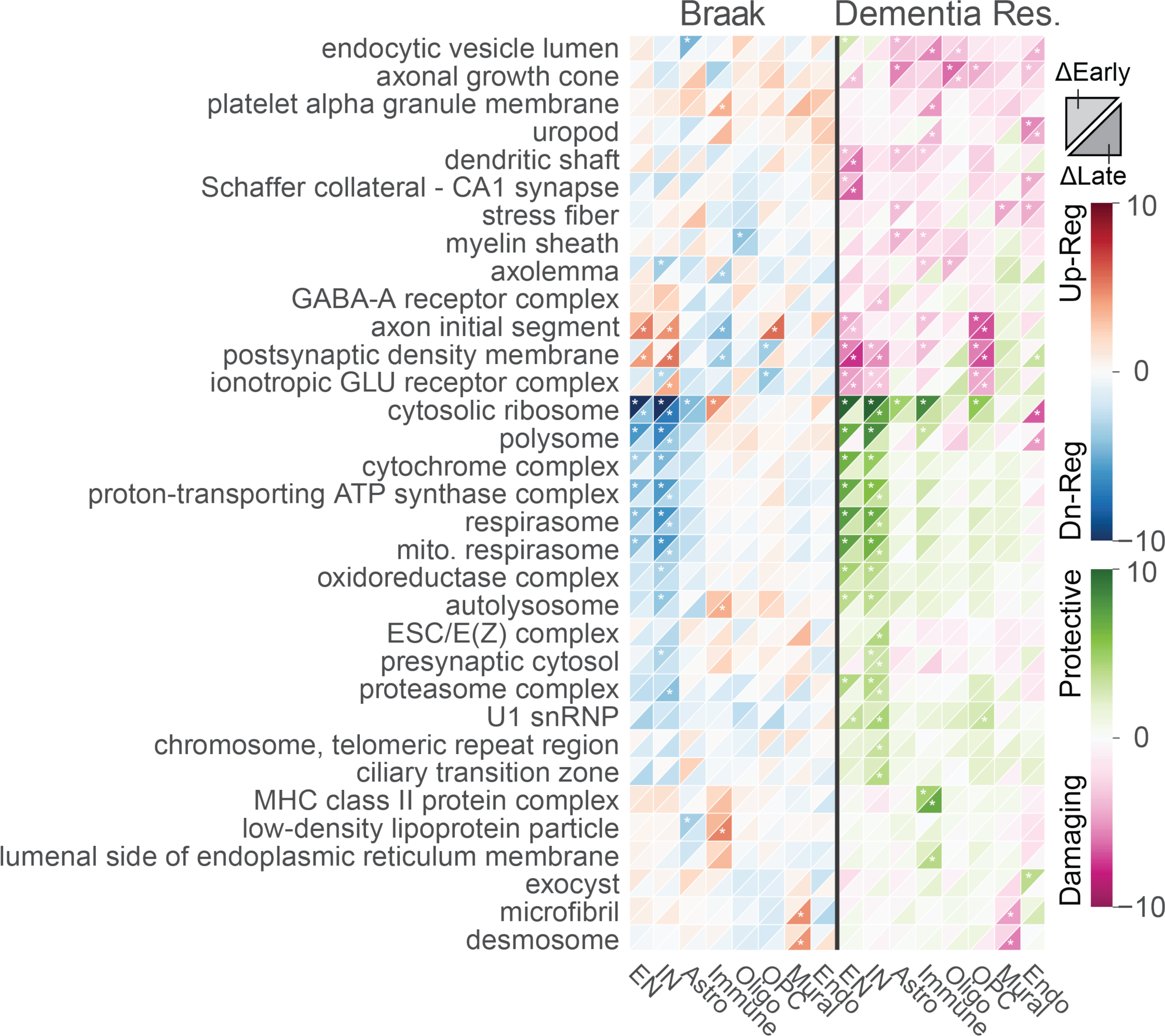
Pathway enrichment using the top GO Cellular Components pathways. Upper left triangles indicate the early phases, and lower right triangles indicate the late phases of the AD trajectory predicted by Braak and dementia resilience. Hue indicates the z-score, and stars indicate FDR < 0.05.

**Supplementary Fig. 11.**
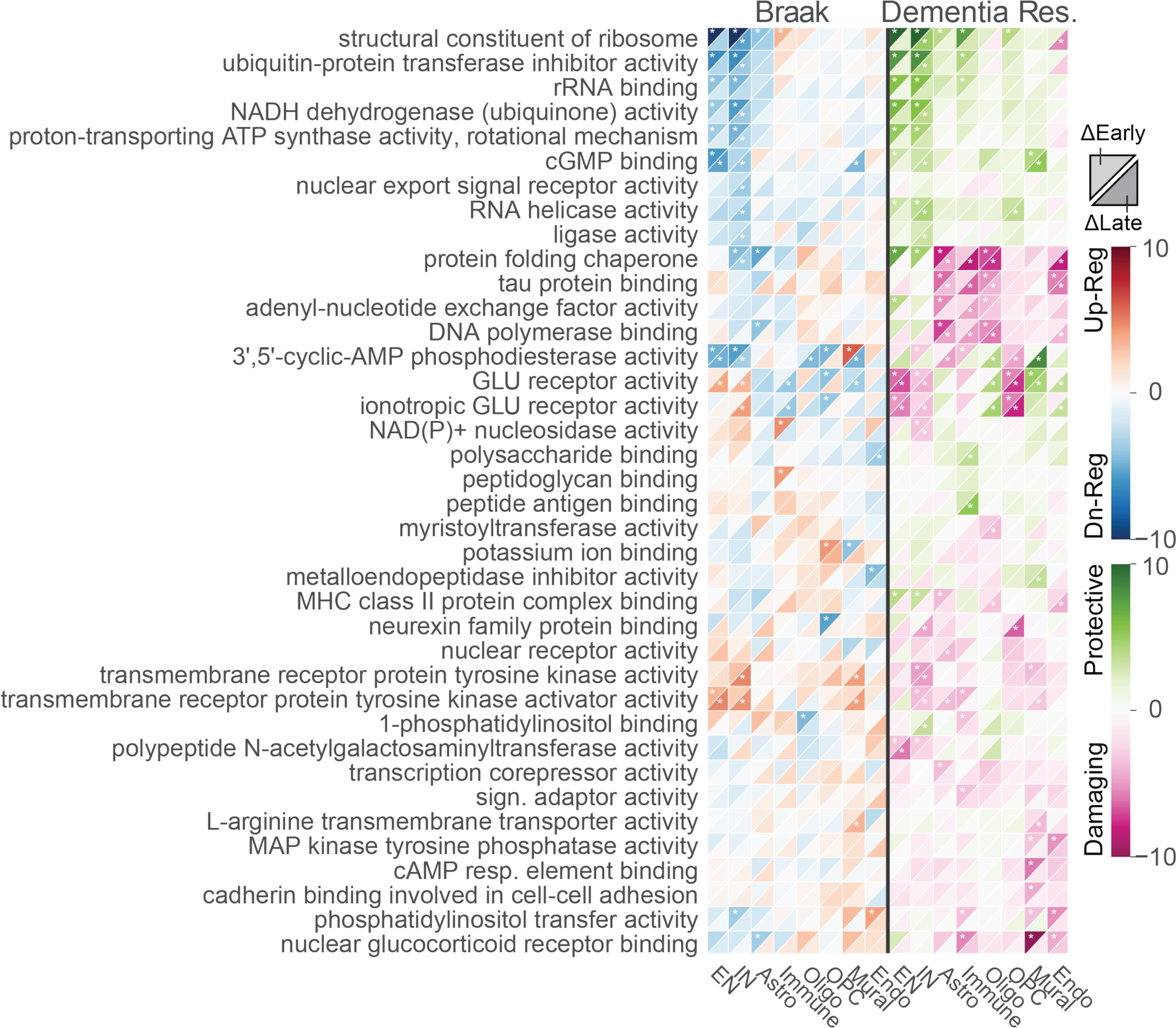
Pathway enrichment using the top GO Molecular Function pathways. Upper left triangles indicate the early phases, and lower right triangles indicate the late phases of the AD trajectory predicted by Braak and dementia resilience. Hue indicates the z-score, and stars indicate FDR < 0.05.

**Supplementary Fig. 12.**
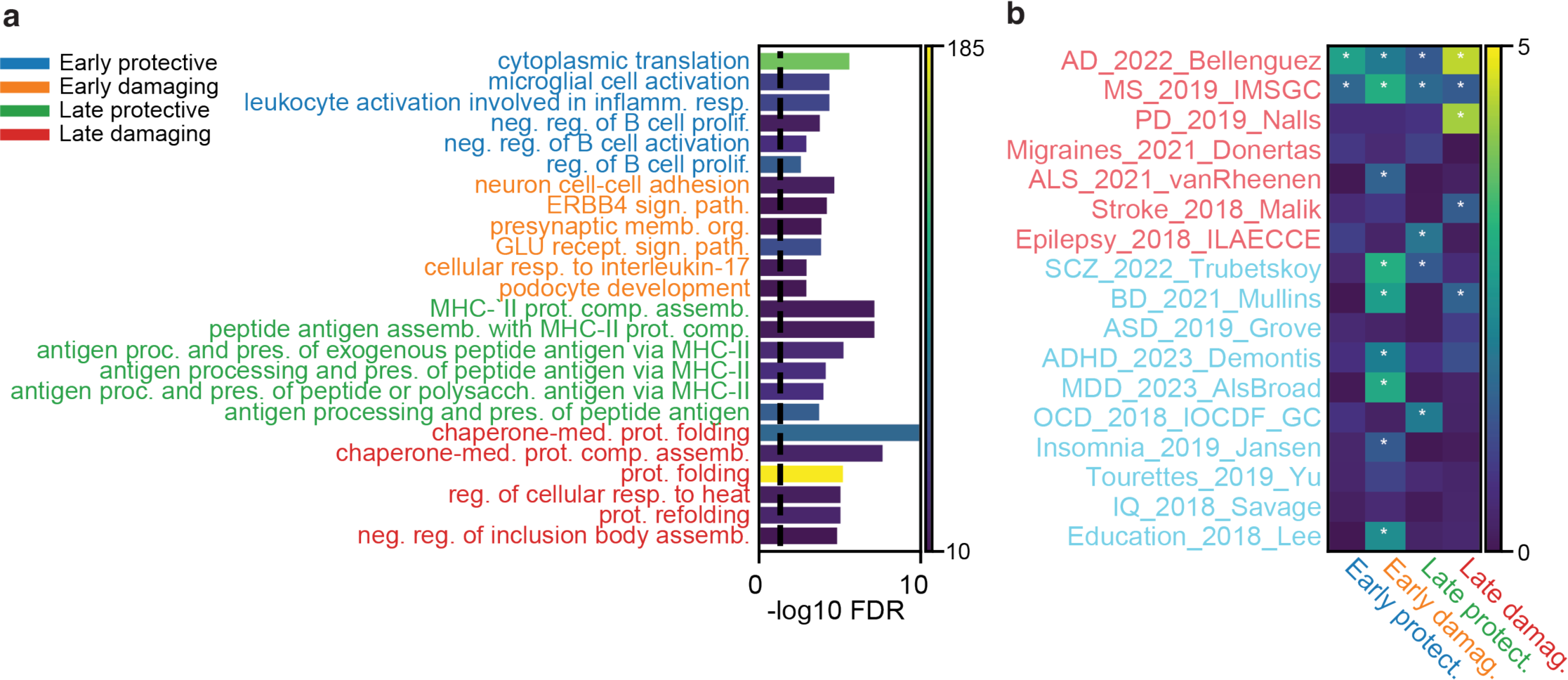
GO BP pathways and GWAS enrichment results for dementia resilience. **(a)** Top early protective pathways (i.e. resilience against dementia, blue), early damaging (i.e. associated with dementia, orange), late protective (green) and late damaging (red). Hue indicates the number of genes in the pathway. Negative log FDR clipped at 10. **(b)** Enrichment of heritability estimates (MAGMA) for each disease trajectory module. Text color indicates neurological traits (light red) and psychiatric traits (cyan). Hue indicates negative log10(FDR), asterisk indicates FDR < 0.05. Negative log FDR clipped at 5.

**Supplementary Fig. 13.**
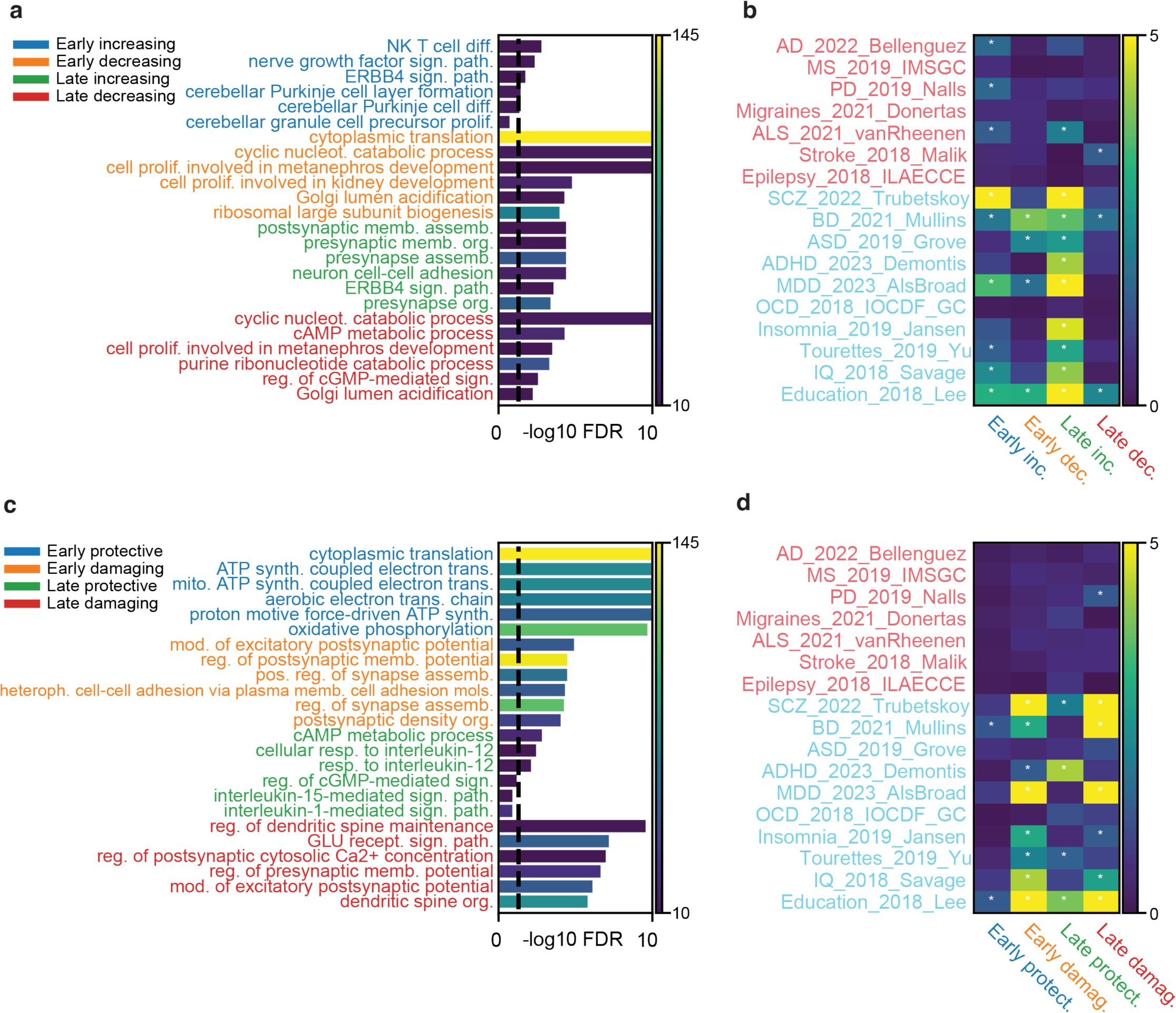
GO BP pathways and GWAS enrichment results for the EN cell class based on their response to Braak (a,b) and dementia resilience (c,d). **(a)** Pathway enrichment of genes with early increasing response to increasing tau proteinopathy (blue), early decreasing (orange), late increasing (green) and late decreasing (red). Hue indicates the number of genes in the pathway. Negative log FDR clipped at 10. **(b)** Enrichment of heritability estimates (MAGMA) for each disease trajectory module. Text color indicates neurological traits (light red) and psychiatric traits (cyan). Hue indicates negative log10(FDR), asterisk indicates FDR < 0.05. Negative log FDR clipped at 5. **(c)** Pathway enrichment of genes with early protective resilience against dementia (blue), early damaging associated with dementia (orange), late protective (green), and late damaging (red). **(d)** Enrichment of heritability estimates (MAGMA) for each disease trajectory module.

**Supplementary Fig. 14.**
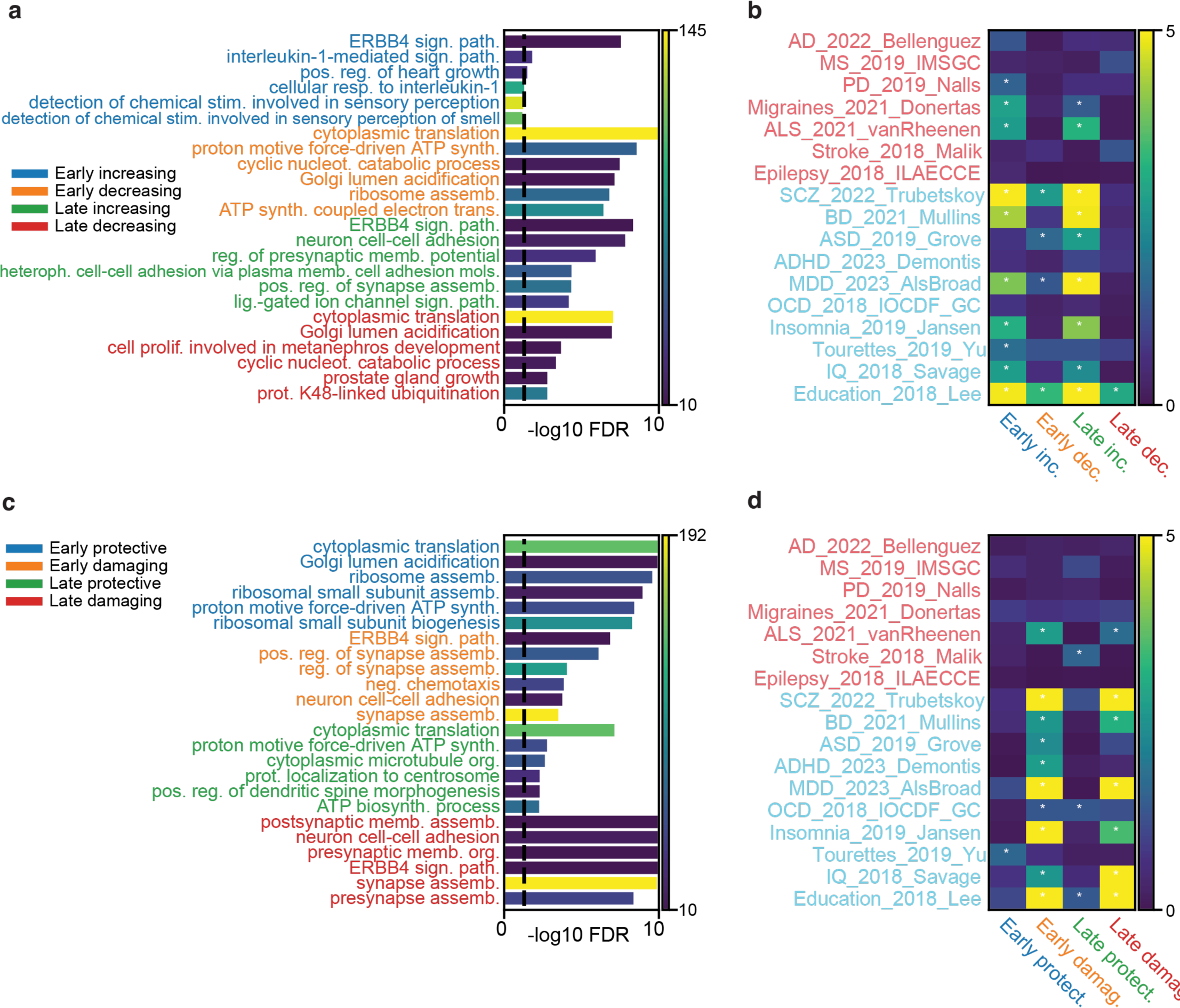
GO BP pathways and GWAS enrichment results for the IN cell class based on their response to Braak (a,b) and dementia resilience (c,d). **(a)** Pathway enrichment of genes with early increasing response to increasing tau proteinopathy (blue), early decreasing (orange), late increasing (green) and late decreasing (red). Hue indicates the number of genes in the pathway. Negative log FDR clipped at 10. **(b)** Enrichment of heritability estimates (MAGMA) for each disease trajectory module. Text color indicates neurological traits (light red) and psychiatric traits (cyan). Hue indicates negative log10(FDR), asterisk indicates FDR < 0.05. Negative log FDR clipped at 5. **(c)** Pathway enrichment of genes with early protective resilience against dementia (blue), early damaging associated with dementia (orange), late protective (green), and late damaging (red). **(d)** Enrichment of heritability estimates (MAGMA) for each disease trajectory module.

**Supplementary Fig. 15.**
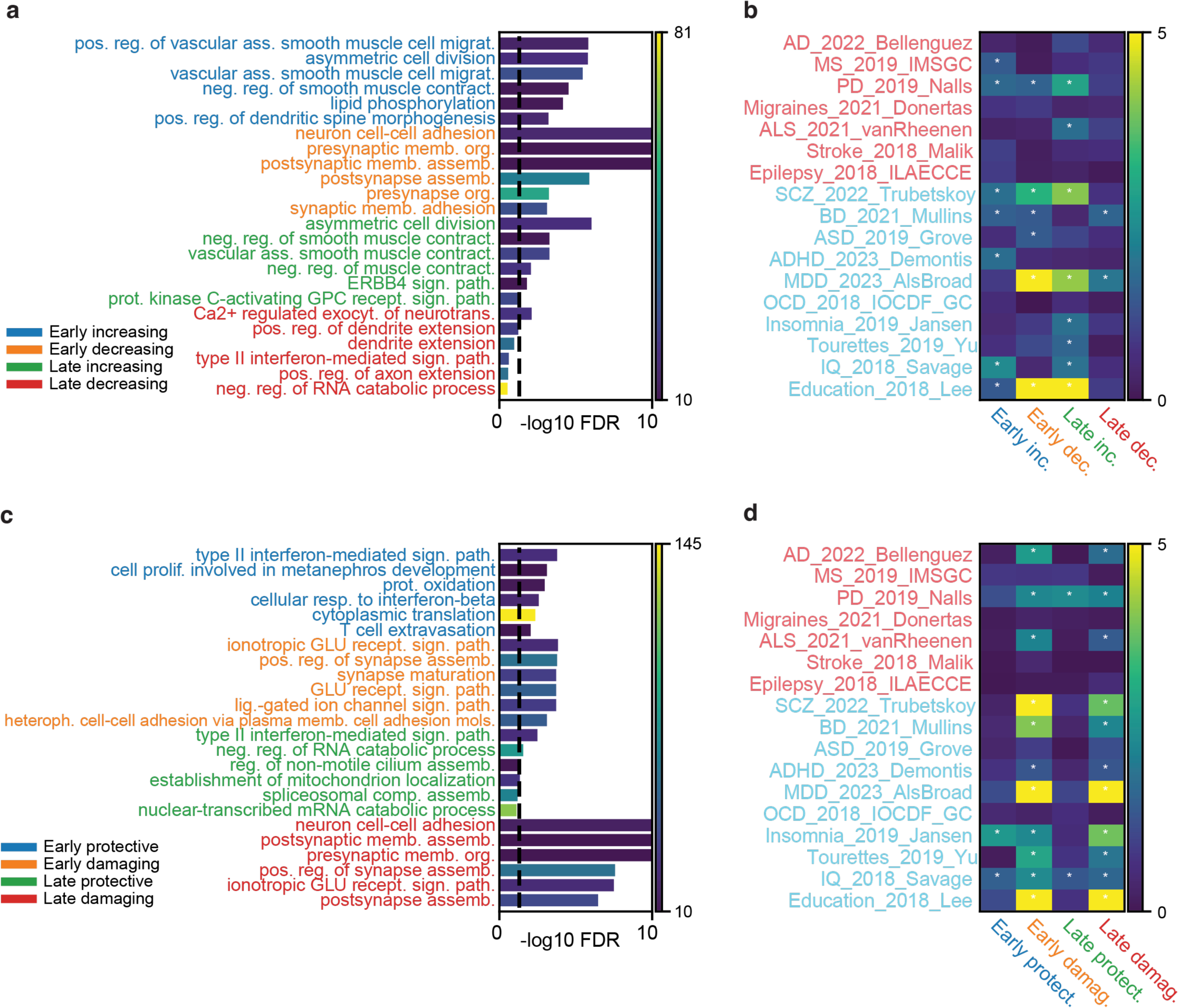
GO BP pathways and GWAS enrichment results for the OPC cell class based on their response to Braak (a,b) and dementia resilience (c,d). **(a)** Pathway enrichment of genes with early increasing response to increasing tau proteinopathy (blue), early decreasing (orange), late increasing (green) and late decreasing (red). Hue indicates the number of genes in the pathway. Negative log FDR clipped at 10. **(b)** Enrichment of heritability estimates (MAGMA) for each disease trajectory module. Text color indicates neurological traits (light red) and psychiatric traits (cyan). Hue indicates negative log10(FDR), asterisk indicates FDR < 0.05. Negative log FDR clipped at 5. **(c)** Pathway enrichment of genes with early protective resilience against dementia (blue), early damaging associated with dementia (orange), late protective (green), and late damaging (red). **(d)** Enrichment of heritability estimates (MAGMA) for each disease trajectory module.

## Supplementary Tables

**Supplementary Table 1.** Clinical and technical metadata of 1,494 donors in the PsychAD cohort. Metadata includes binary contrasts and continuous variables used in the study.

**Supplementary Table 2.** Hierarchical cellular taxonomy of the PsychAD snRNA-seq data. Metadata including parent-child relationships, color hex codes, and number of nuclei per subtype.

**Supplementary Table 3.** Compositional variation analysis across 8 cross-disorder traits.

**Supplementary Table 4.** Differentially expressed genes across 8 cross-disorder traits.

**Supplementary Table 5.** Compositional variation analysis across AD phenotypes.

**Supplementary Table 6.** Differentially expressed genes across AD phenotypes.

**Supplementary Table 7.** The top 50 GO BP pathways for each cell class relative to early/late increases/decreases in predicted Braak staging, and relative to early/late dementia resilience

**Supplementary Table 8.** Gene-set enrichment analysis using GWAS summary data for each cell class. GWAS scores are calculated using the top 250 coding genes with the greatest slopes measured relative to early/late increases/decreases in predicted Braak staging, and the top 250 early/late protective/damaging coding genes.

**Supplementary Table 9.** The top 250 coding genes with the greatest slopes measured relative to early/late increases/decreases in predicted Braak staging, and the top 250 early/late protective/damaging coding genes.

**Supplementary Table 10.** List of gene markers used from Xenium human brain custom panel.

### Supplementary Data

**Supplementary Data 1**. Metadata and model predictions for each cell. Each cell class contains a table that includes the cell barcode, donor information, the cell subclass, and both the actual Braak and dementia status along with the model predictions.

## Notes

### Competing Interest Statement

The authors have declared no competing interest.

### Author Declarations

Ethics committee/IRB of Mount Sinai gave ethical approval for this work. Ethics committee/IRB of James J. Peters Department of Veterans Affairs Medical Center gave ethical approval for this work. Ethics committee/IRB of Rush Alzheimer's Disease Center gave ethical approval for this work. Ethics committee/IRB of National Institute of Mental Health Human Brain Collection Core gave ethical approval for this work.

